# Neuromechanical gait signatures reveal holistic biomechanical responses to walking speed modulation in stroke survivors

**DOI:** 10.1101/2022.12.14.22283438

**Authors:** Michael C. Rosenberg, Taniel S. Winner, Gordon J Berman, Lena H. Ting, Trisha M. Kesar

## Abstract

During gait rehabilitation, determining the walking speed that optimizes an individual’s biomechanical gait quality is challenging because discrete biomechanical variables used to quantify gait quality change differentially with speed. We recently developed *gait signatures* that may provide a holistic representation of biomechanical gait quality by implicitly modeling the neuromechanical dynamics of walking. Here, we characterized speed-induced changes in post-stroke gait signatures and their relationship to 14 discrete biomechanical variables in 19 stroke survivors and 5 able-bodied adults walking at 6 speeds. With increasing speed, post-stroke gait signatures became more like able-bodied gait signatures, suggesting improved gait quality. Inter-individual differences in the direction that post-stroke gait signatures changed with speed relative to able-bodied signatures were correlated with walking function (e.g., walking speed; r^2^=0.53) and discrete biomechanical variables (e.g., paretic-leg propulsion; r^2^*=*0.57). These findings suggest that *how* post-stroke gait signatures change with speed relative to able-bodied signatures reflects neuromechanical constraints that also impact biomechanical gait quality. Across speeds, gait signatures captured holistic biomechanical gait quality, balancing tradeoffs between improved paretic-leg biomechanics and worsened inter-limb asymmetry and gait compensations (r^2^=0.77). Gait signatures may, therefore, be useful in holistic assessment of inter-and intra-individual differences in biomechanical gait quality, potentially informing rehabilitation personalization.

## INTRODUCTION

Selecting individual-specific gait rehabilitation treatment parameters to improve walking speed while maximizing the overall quality of gait biomechanics remains challenging. Walking speed is reduced following neurological injuries or neurodegenerative disorders such as a stroke or Parkinson’s disease^1–3^. Individuals with neuropathologies often exhibit slower walking speeds, limiting community activity^4,5^. Increasing an individual’s walking speed is, therefore, a popular goal of gait rehabilitation^2,6^. However, increasing walking speed during rehabilitation may be accompanied by breakdowns in biomechanical gait quality, potentially reducing gait efficiency and overall musculoskeletal health^7–10^. Gait quality can be measured using spatiotemporal, kinematic, and/or kinetic variables of the affected leg(s), inter-limb asymmetry, and gait compensations^9,11–14^. However, the speed that maximizes gait quality differs across biomechanical metrics and varies across individuals^7,9,12^. Gait rehabilitation designed to increase walking speed without sacrificing biomechanical gait quality may, therefore, benefit from a more holistic approach to understanding how gait biomechanics change with speed. We posit that the complexity of post-stroke gait deficits and challenges quantifying how walking speed impacts gait biomechanics necessitate the application of novel methodological frameworks to understand how modulating gait speed holistically impacts biomechanical gait quality.

Studies in stroke survivors reveal that different discrete biomechanical variables may produce conflicting conclusions as to whether walking faster improves or degrades biomechanical gait quality. During overground or treadmill-based post-stroke gait retraining, walking speed should ideally be modulated to maximize training intensity, while also selecting an “optimal” fast speed that an individual can maintain without degradations in biomechanical gait quality^15^. However, changes in post-stroke gait biomechanics with increasing walking speed are individual-specific; speed-induced changes in both the magnitude of paretic-leg biomechanics and inter-limb and inter-joint coordination patterns vary between individuals^11,12,16^. Within-individuals, post-stroke biomechanical variables also change differentially with speed; stroke survivors often improve paretic-leg biomechanics at faster walking speeds, with more variable changes in inter-limb asymmetry and gait compensations (e.g., paretic-leg circumduction)^7,8,12,16,17^. For example, during the stance phase of the paretic leg, anterior ground reaction forces (AGRFs), trailing limb angle (TLA), and ankle and hip moments and powers typically improve (i.e., increase) with faster walking speeds^8,18,19^. Conversely, swing-phase paretic-leg compensations exhibit smaller and more variable changes with increasing walking speed, though the functional importance of these small gait deviations remains unclear^7,9^. Inter-limb asymmetry, defined here as the difference in paretic and non-paretic legs biomechanics, also changes heterogeneously with increasing speed^12,13^.

Neuromechanical *gait signatures* characterize individual differences in able-bodied (AB) and post-stroke gait dynamics across walking speeds, but whether they holistically encode inter-individual differences in post-stroke biomechanical gait quality across walking speeds is unclear^20,21^. We recently developed the gait signatures modeling framework to capture and compare gait neuromechanics (i.e., the neural and biomechanical constraints governing gait) across individuals and walking speeds. To capture inter-and intra-individual differences in the constraints governing the time-evolution of joint kinematics (i.e., gait dynamics), the gait signatures framework uses an artificial recurrent neural network (RNN) to encode gait dynamics across individuals and speeds. Individual-and trial-specific gait signatures are constructed from the RNN’s latent states and can be compared between individuals and speeds. Gait signatures consist of multidimensional stride-averaged waveforms of the RNN’s latent states, which represents how an individual’s neuromechanical dynamics evolve over the gait cycle for each walking speed. The similarity of gait signatures can be quantified as the Euclidean distance between waveforms for two individuals or walking speeds^20^. In an initial study of 7 stroke survivors, only some participants’—predominantly faster-walking participants’—gait signatures became more similar to those of able-bodied (AB) adults with increasing walking speed^20^. Gait signatures identified individuals across walking speeds, suggesting that they reflect inter-individual differences in gait dynamics. However, whether gait signatures encode holistic individual differences in post-stroke gait biomechanics across walking speeds has not been investigated. We recently showed that in AB young adults walking at a wide range of speeds, gait signatures changed in a similar direction across all participants and changed linearly with speed for 11 of 17 participants^21^. While the magnitude of within-individual changes in gait signatures with walking speed was correlated with changes in step length and cadence. However, this prior study did not analyze relationships between gait signatures and other biomechanical variables used to quantify post-stroke gait quality.

Which features of gait signatures best encode individual differences in post-stroke biomechanical gait quality is also unclear. Because common post-stroke biomechanical variables (e.g., AGRF, TLA) are computed at discrete timepoints within the gait cycle, differential effects of speed on post-stroke gait signatures on the stance versus swing phases of the gait cycle may impact the relationship between gait signatures and biomechanical variables^7–9,19^. Further, different gait signatures metrics may encode different information about post-stroke biomechanical gait quality. Consistent with previously observed increases in both post-stroke gait signature similarity to AB adults^20^ and improvements in some biomechanical variables^8^ with walking speed, as well as the framework of restitution versus compensation as a gait recovery mechanism^22^, we expect that greater similarity of post-stroke gait signatures to AB gait signatures will encode better biomechanical gait quality. However, our recent finding in AB adults gait signatures change with speed in a similar direction across individuals suggests that *how* gait signatures change with speed may better reflect the neural and biomechanical constraints governing speed-induced changes in post-stroke gait biomechanics^11,21,23–25^. Additionally, scalar metrics of how much gait signatures change with speed may be too coarse to encode holistic biomechanical gait quality. Metrics that leverage the multidimensional nature of gait signatures and gait biomechanics may better encode holistic biomechanical gait quality^7,9,16^.

Here, we characterized whether post-stroke gait signatures holistically encode biomechanical gait quality across walking speeds. We hypothesized that, because gait signatures encode individual differences in gait dynamics, gait signatures will reflect holistic individual differences in post-stroke gait biomechanics across walking speeds. Based on our prior findings^21^, we expected that the similarity of post-stroke gait signatures to an AB reference gait signature and the similarity of the *direction* in which post-stroke gait signatures changed with speed compared to an AB reference gait signature would capture different aspects of biomechanical gait quality. We predicted that: (1) Greater similarity of baseline (i.e., self-selected (SS) speed) post-stroke gait signatures to an AB *reference* (i.e., average) gait signature would be associated with greater quality of individual biomechanical variables; (2) Greater similarity in the direction of speed-induced changes in post-stroke gait signatures relative to that of AB reference gait signatures would be associated with greater improvements in the quality of individual biomechanical variables at faster walking speeds; (3) Across speeds, greater similarity of post-stroke gait signatures to the AB reference gait signature would be associated with greater holistic biomechanical gait quality (i.e., across all biomechanical variables). To further assess gait signatures as a holistic representation of biomechanical gait quality, we show that gait signatures capture individual differences in biomechanical gait quality more holistically than walking speed alone.

## RESULTS

### Recurrent-neural-network-based gait signatures identified inter-and intra-individual differences in gait dynamics from gait kinematics in AB adults and stroke survivors across 6 walking speeds

Following the procedure of Winner and colleagues (2023), gait signatures were computed from sagittal-plane joint kinematics in 5 AB adults and 20 stroke survivors walking on a treadmill at six speeds ranging from their SS to fastest safe speed (Figure 1A)^20^. A single RNN layer with long short-term memory (LSTM) activation units (Figure 1B) was trained on timeseries data to predict sagittal-plane hip, knee, and ankle kinematics one time-step (10ms) in the future using the same kinematic variables at the current time-step as inputs. To enable comparisons across participants and walking speeds, a single model was trained on all participants’ data. Gait signatures were computed by projecting the LSTM’s trial-specific, time-varying latent states into a low-dimensional basis using principal components analysis (PCA) and phase-averaging the principal components (PCs) (Figure 1B & C)^26^.

**Figure 1:**
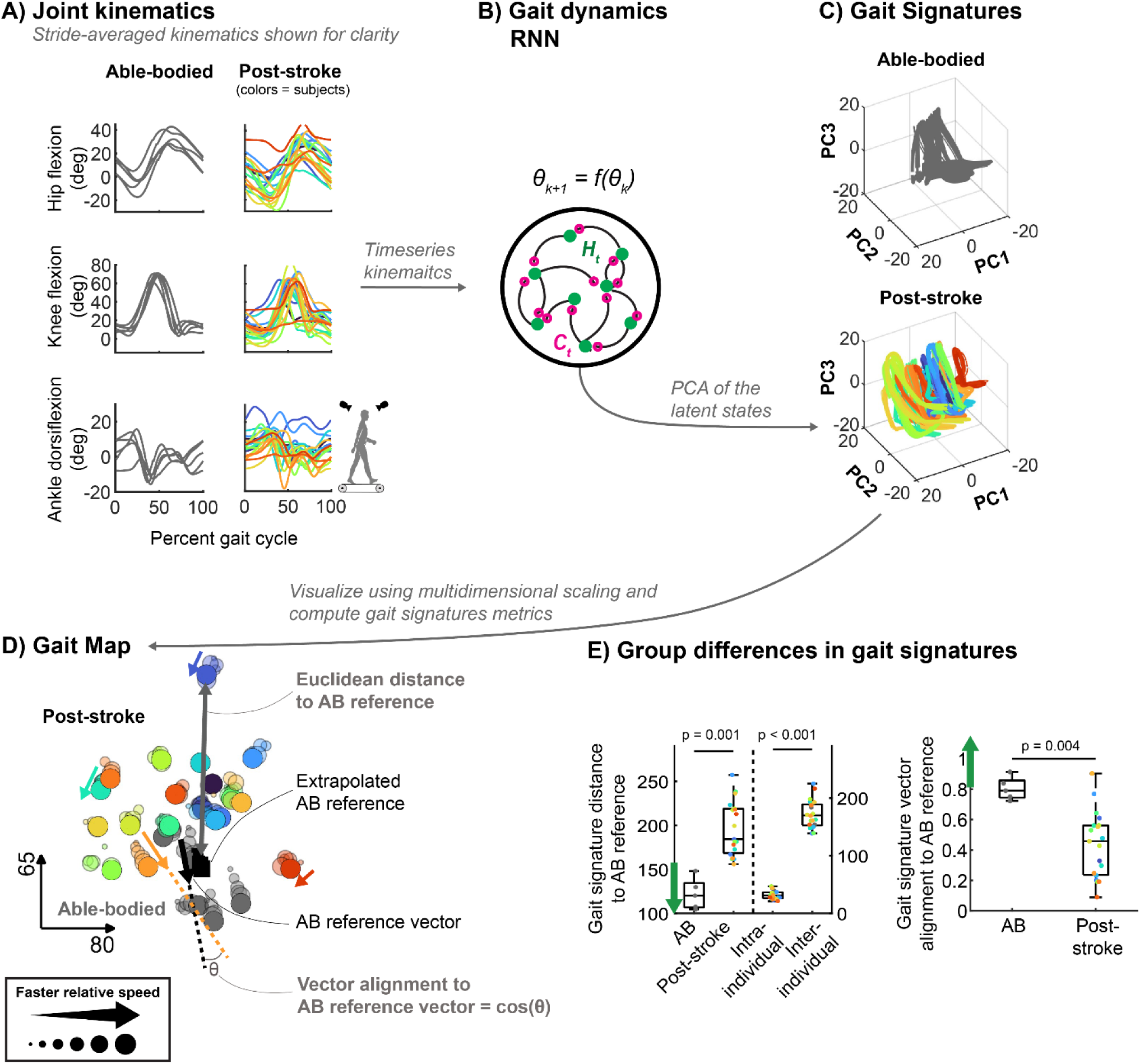
Overview of the Gait Signatures pipeline. **A)** Examples of stride-averaged kinematics at each participant’s SS speed. Stride-averaged kinematics are shown for clarity only; continuous timeseries kinematics (15s; 1500 samples per trial) across all participants and walking speeds were used to train the gait dynamics recurrent neural network (RNN). Each participant walked at 6 speeds, ranging from their self-selected to their fastest comfortable speed. Only one speed per participant is shown for clarity. Gray lines denote able-bodied adults. Each post-stroke participant is denoted by a different color that is maintained across figures. **B)** The concatenated data were used to train a single RNN. The dominant patterns in the RNN’s latent states were extracted using principal components analysis (PCA). The first 6 PCs were used to construct participant-and trial-specific *gait signatures*. **C)** The first three PCs are shown on separate plots for able-bodied (top) and post-stroke participants (bottom). **D)** We used Multidimensional Scaling to produce a Gait Map representing the relative similarity of gait signatures between trials (dots; larger dots denote faster speed relative to the participant’s self-selected speed) and participants (colors denote stroke survivors; gray denotes able-bodied adults). The black square denotes the AB reference gait signature. The gray double-headed arrow shows an example Euclidean distance to the AB reference gait signature. Colored single-headed arrows show sample participants’ gait signature vectors (i.e., the direction of change in gait signatures between the SS and fastest speeds). The black single-headed arrow represents the corresponding vector for the AB reference gait signatures at the SS and fastest speeds. Gait signature vector alignment was defined as the projection of post-stroke vectors onto the AB reference vector, or the cosine of the angle between the vectors. **E; left)** Group, inter-and intra-individual gait signature distances to the AB reference. **E; right)** Group differences in gait signature vector alignment to the AB reference. Green arrows denote the direction of more AB-like gait signatures and vectors.

The first 6 PCs of the latent states accounted for 92% of the variance in the latent states and were included in subsequent analyses (*Supplemental Figure S1*). One participant was omitted from subsequent analyses due to data corruption during the fastest-speed trial.

All AB participants’ gait signatures had similar shape and amplitude, whereas post-stroke participants’ gait signatures exhibited larger inter-individual differences in shape and amplitude when visualizing the first 3 components (PCs) of the gait signatures for each trial (Figure 1C gray vs colors; traces represent single trials). The 2D *gait map* in Figure 1D represents the relative similarity of each gait signature and was computed using Multidimensional Scaling.

Post-stroke gait signatures were less similar to the AB reference gait signature than were AB gait signatures. We defined the similarity of gait signatures across walking speeds as the *Euclidean distance* of each gait signature to the AB reference gait signature. More-similar pairs of gait signatures had smaller Euclidean distances (Figure 1D; gray double-headed arrow represents the distance to the AB reference gait signature). The Euclidean distances of SS-speed post-stroke gait signatures to the AB reference gait signature were greater than those of AB adults (p = 0.001; Cohen’s *d* = 2.6; Figure 1E; left). Across participants, gait signatures were also individual-specific: within-participant Euclidean distances in gait signatures (i.e., between walking speeds) were smaller than between-participant differences (p = 3.1e-9; Cohen’s *d* = 8.0; Figure 1E; left).

Post-stroke gait signatures changed in different directions compared to those of AB gait signatures. We defined this directional change using the *gait signature vector alignment* relative to an AB reference gait signature vector. Gait signature vector alignment represents the similarity of the direction that each participant’s gait signature changed with speed, relative to that of the average AB gait signature vector (Figure 1D; angle of colored arrows relative to the black single-headed arrow). We computed vector alignment as the inner product of a unit vector between each participant’s SS and fastest-speed gait signatures (Figure 1D; colored arrows denote directions of change with speed) and a corresponding unit vector computed for the AB reference gait signature. Greater vector alignment indicates that gait signatures change with speed in a more-similar direction to that of the AB reference gait signature.

Post-stroke vector alignment was lower than that of AB participants (p = 0.004; Cohen’s *d* = 1.8; Figure 1E; right).

### Gait signature vector alignment, but not Euclidean distances to the AB reference gait signature, was correlated with clinical measures of walking function

Across post-stroke participants, SS-speed gait signature Euclidean distances to the AB reference gait signature were only marginally correlated with treadmill-based SS walking speeds (r^2^ = 0.22; p = 0.045; α_Sidak_ = 0.010) and lower-extremity Fugl-Meyer (FM-LE) scores (r^2^ = 0.31; p = 0.014; Figure 2A), and were not correlated with overground SS speed or speed range (p > 0.188). In contrast, better post-stroke gait signature vector alignment was correlated with faster treadmill-based SS speeds (r^2^ = 0.53; p = 4.0e-4; Figure 2B), faster overground SS speeds (r^2^ = 0.53; p = 4.2e-4), and a greater range of walking speeds (r^2^ = 0.65; p = 3.0e-5).

**Figure 2:**
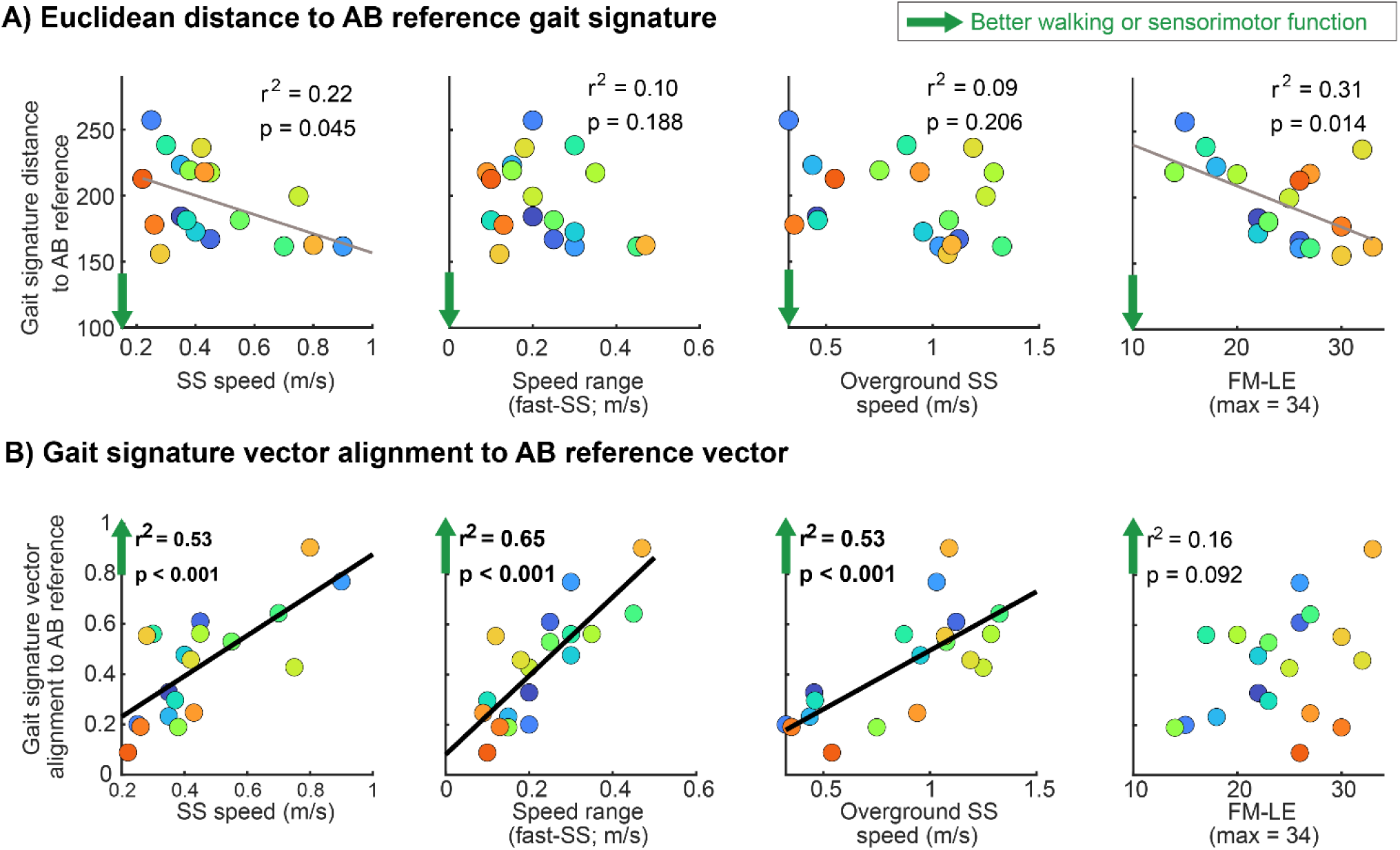
Correlations between SS-speed gait signatures and clinical measures of walking and motor function. Colored dots denote post-stroke participants and gray dots denote AB participants. Colors are preserved across panels. Green arrows denote the director of more AB-like gait signatures (top) and vectors (bottom). The clinical metrics (columns) are: treadmill-walking SS-speed (left), treadmill walking speed range (fastest vs. SS-speed; center-left), overground SS speed (center-right), and lower-extremity Fugl-Meyer score (FM-LE; right). **A)** Clinical metrics regressed against the Euclidean distances between post-stroke gait signatures and the AB reference gait signature. **B)** Clinical metrics regressed against post-stroke participants’ gait signature vector alignment to the AB reference vector. Boldfaced r^2^ and p-values indicates significant correlations after correction for multiple comparisons (α_Sidak_ = 0.010).

### Post-stroke gait signatures became more similar to AB gait signatures at faster walking speeds, and typically did so during both the stance and swing phases of the paretic leg

Compared to the SS walking speed, post-stroke gait signatures at the fastest speed were more-similar to the AB reference gait signature over the entire gait cycle and in the paretic-leg stance and swing phases of the gait cycle (Figure 3A; all p < 1.9e-4). Post-stroke gait signature Euclidean distances to the AB reference were minimized at faster speeds more frequently than swing-phase gait signatures (Figure 3B; yellow dots). Full-gait-cycle and stance-phase gait signature Euclidean distances were minimized at the fastest speed in 58% of participants (Figure 3C; black and orange, respectively), while swing-phase Euclidean distances were minimized at the fastest speed in 42% of participants (Figure 3D; blue). Slower-walking stroke survivors’ gait signatures changed less with increasing walking speed than those of faster-walking stroke survivors in both the stance and swing phases of the gait cycle (Figure 3D; AB gait signatures shown for reference). Faster-walking stroke survivors (pink; SS speed > 0.40 m/s) tended to change gait signatures more than slower-walking stroke survivors (purple; SS speed ≤ 0.40 m/s), who exhibited smaller changes in gait signatures with speed and smaller speed ranges. Individual-specific correlations between speed and gait signature Euclidean distances can be found in *Supplemental Figure S3*.

**Figure 3:**
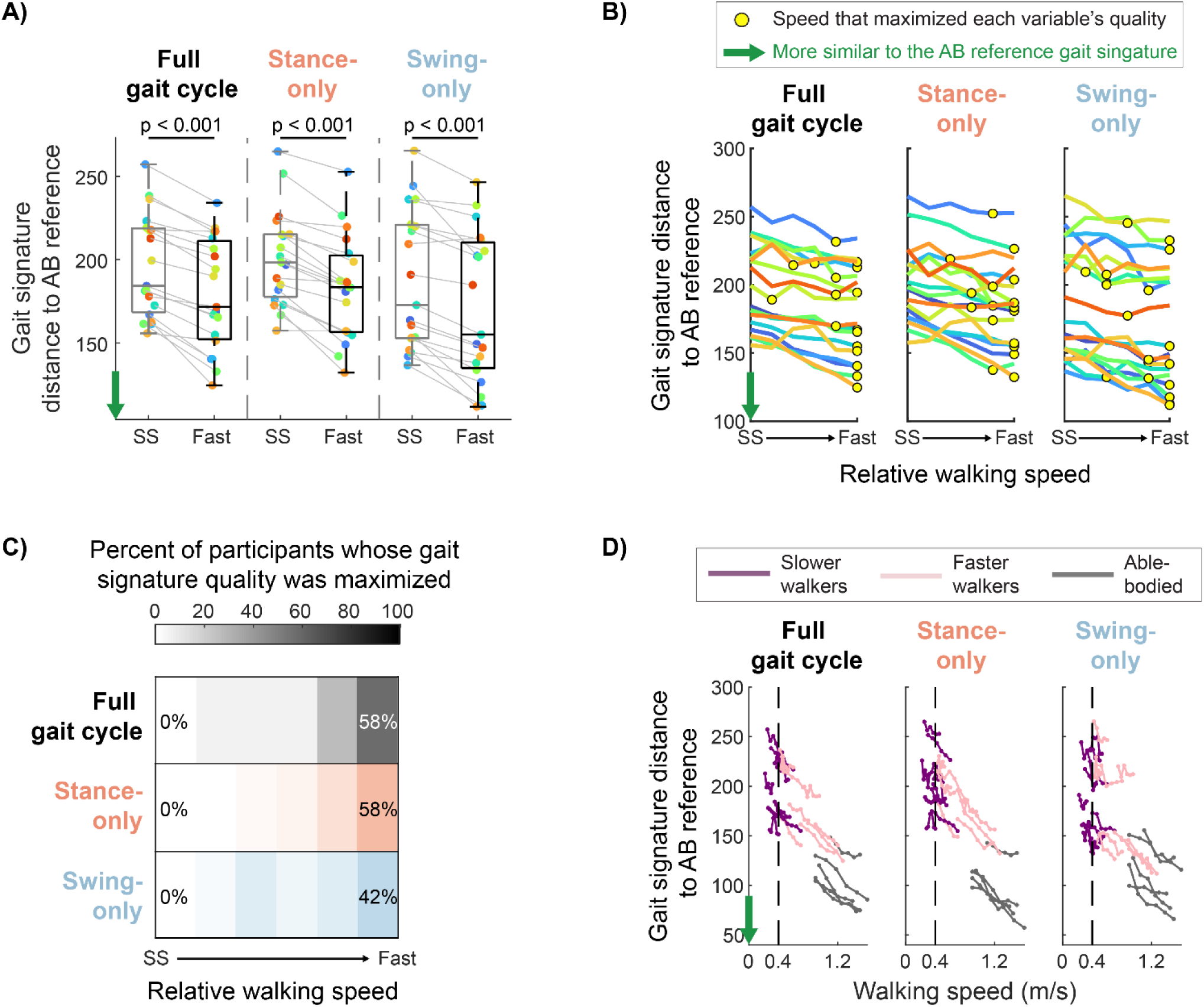
Differential effects of walking speed on gait signatures. Green arrows denote the direction of more AB-like gait signatures. **A)** Gait signature distances to the AB reference gait signature at the SS (gray) and fastest (black) walking speeds. Distances were computed over the entire gait cycle, only the paretic-leg stance phase of the gait cycle (stance-only), and only the paretic-leg swing-phase of the gait cycle (swing-only). Colored dots denote participants.

### Post-stroke discrete biomechanical variables and gait signatures changed differentially with speed within-and between-participants, but inter-limb asymmetry and gait compensations did not reliably improve at faster speeds

Across participants, paretic-leg magnitude variables tended to improve with increasing walking speed; biomechanical magnitude variables improved at the fastest speed in 58-79% of participants (Figure 4A, green heatmap corresponds to the number of yellow dots at each speed in Figure 4B; yellow dots represent the speed that maximally improved each biomechanical variable). Inter-limb asymmetry variables were most improved (i.e., asymmetry was minimized) in only 0-26% of participants at the fastest speed (Figure 4A, orange heatmap) and 16-53% of participants at the SS speed. Gait compensation variables were most improved (i.e., compensations were minimized) in only 11-21% of participants at the fastest speed and 21-42% of participants at the SS speed (Figure 4A, purple heatmap). While paretic-leg biomechanical variables were not maximally improved at the fastest walking speed across participants, paretic-leg AGRFs, TLA, hip power, and ankle power and moments tended to improve (i.e., increase) with increasing speed (Figure 4B; left; green arrows denote direction of improved biomechanical quality). Conversely, inter-limb asymmetry and compensation variables changed heterogeneously with walking speed and often worsened (i.e., increased) with increasing walking speed (Figure 4B; middle and right).

**Figure 4:**
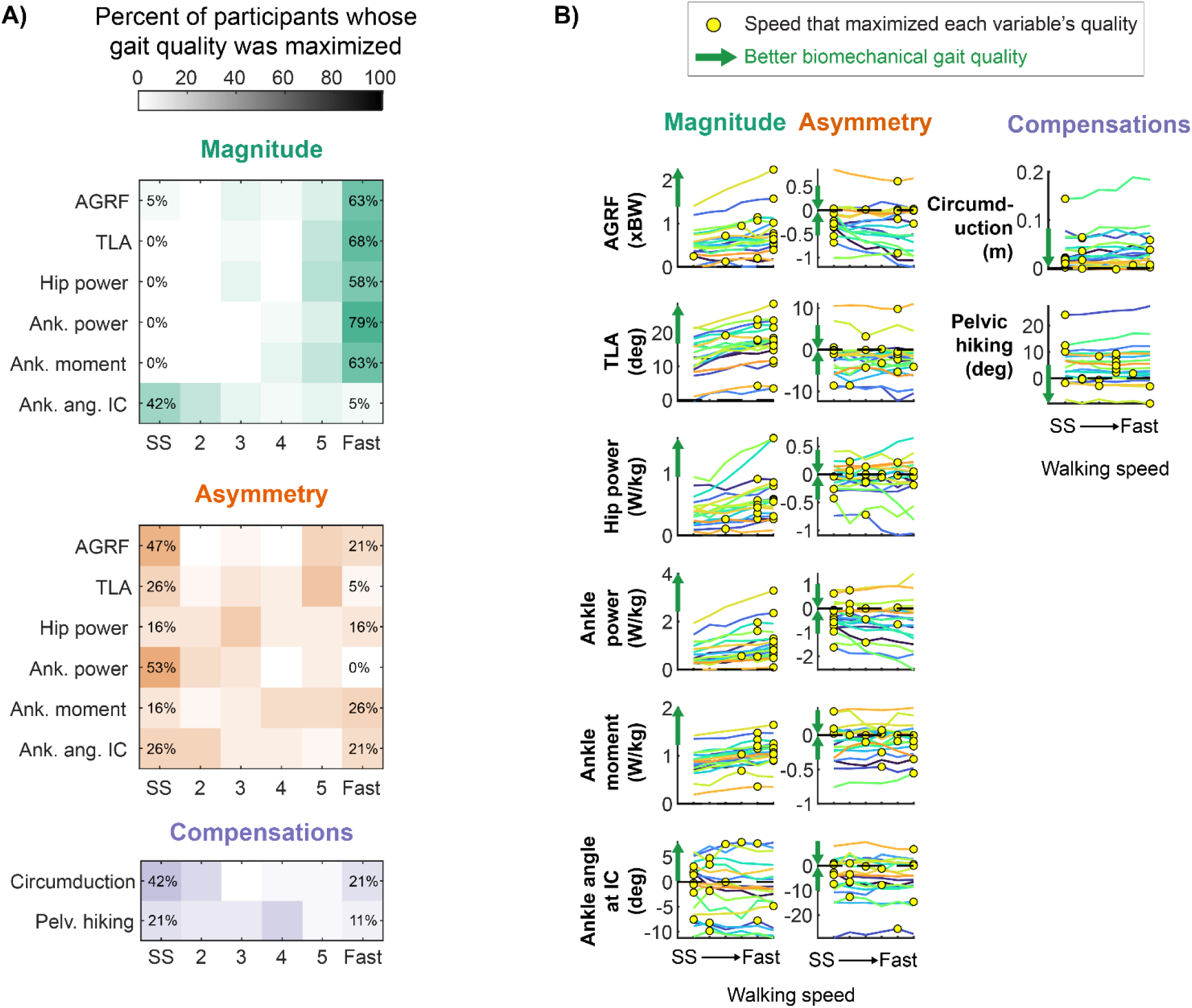
Individual-specific effects of walking speed on discrete biomechanical variables in stroke survivors. **A)** Changes in the magnitude of paretic-leg biomechanics (top) and inter-limb asymmetry (bottom; paretic - non-paretic) for each participant. Colors denote participants. Speeds are shown as speed indices between the self-selected (SS) and fastest (Fast) speed for each participant. Yellow dots denote the speed the maximizes the quality of each variable for each participant. These plots are analogous to those shown for gait signatures in Figure 3B. For each variable, green arrows denote the direction of improved biomechanical quality**. B)** Heatmaps showing the percentage of participants for whom each speed maximized the quality of each biomechanical variable. Separate heatmaps are shown for paretic-leg magnitude (green), inter-limb asymmetry (orange), and compensation (purple) variables. All heatmaps span 0-100% of participants. Abbreviations: AGRF = anterior ground reaction force; Ank. Ang. IC = ankle angle at initial contact; Ank. mom. = peak ankle plantarflexor moment; Ank. pwr. = peak ankle plantarflexor power; Circ. = circumduction; FM-LE = lower-extremity Fugl-Meyer score; Hip pwr. = Peak hip power; Pelv. Hiking = pelvic hiking; TLA = trailing limb angle.

### Speed-induced changes in post-stroke gait signatures, not SS-speed Euclidean distances to the AB reference gait signature, were correlated with differences in some paretic-leg discrete biomechanical variables

At SS walking speeds, post-stroke participants’ gait signature Euclidean distances to the AB reference were not correlated with any discrete biomechanical variables (r^2^ ≤ 0.30; p > 0.015; α_Sidak_ = 9.8e-4; Figure 5A). In contrast, greater post-stroke gait signature vector alignment was correlated with greater SS-speed paretic-leg AGRFs (r^2^ = 0.57 p = 1.7e-4), but was not correlated with inter-limb asymmetry or compensation variables (Figure 5B). Between the SS and fastest walking speeds, greater speed-induced changes in post-stroke gait signature Euclidean distances toward the AB reference were correlated with increases in peak ankle power (r^2^ = 0.67; p = 2.0e-5; α_Sidak_ = 9.8e-4; Figure 5C; left column) and marginally with increases in paretic-leg AGRFs (r^2^ = 0.45; p = 0.002), but not with changes in inter-limb asymmetry or compensatory biomechanical variables. Similarly, greater gait signature vector alignment was correlated with speed-induced increases in paretic-leg AGRF (r^2^ = 0.59; p = 1.1e-4) and peak ankle power (r^2^ = 0.69; p = 1.0e-5; Figure 5D; left column), but not with increases in inter-limb asymmetry or compensatory biomechanical variables. Correlations between post-stroke Euclidean distances to the AB reference, gait signature vector alignment, and all discrete biomechanical variables can be found in *Supplemental Figures S4 & S5*.

**Figure 5:**
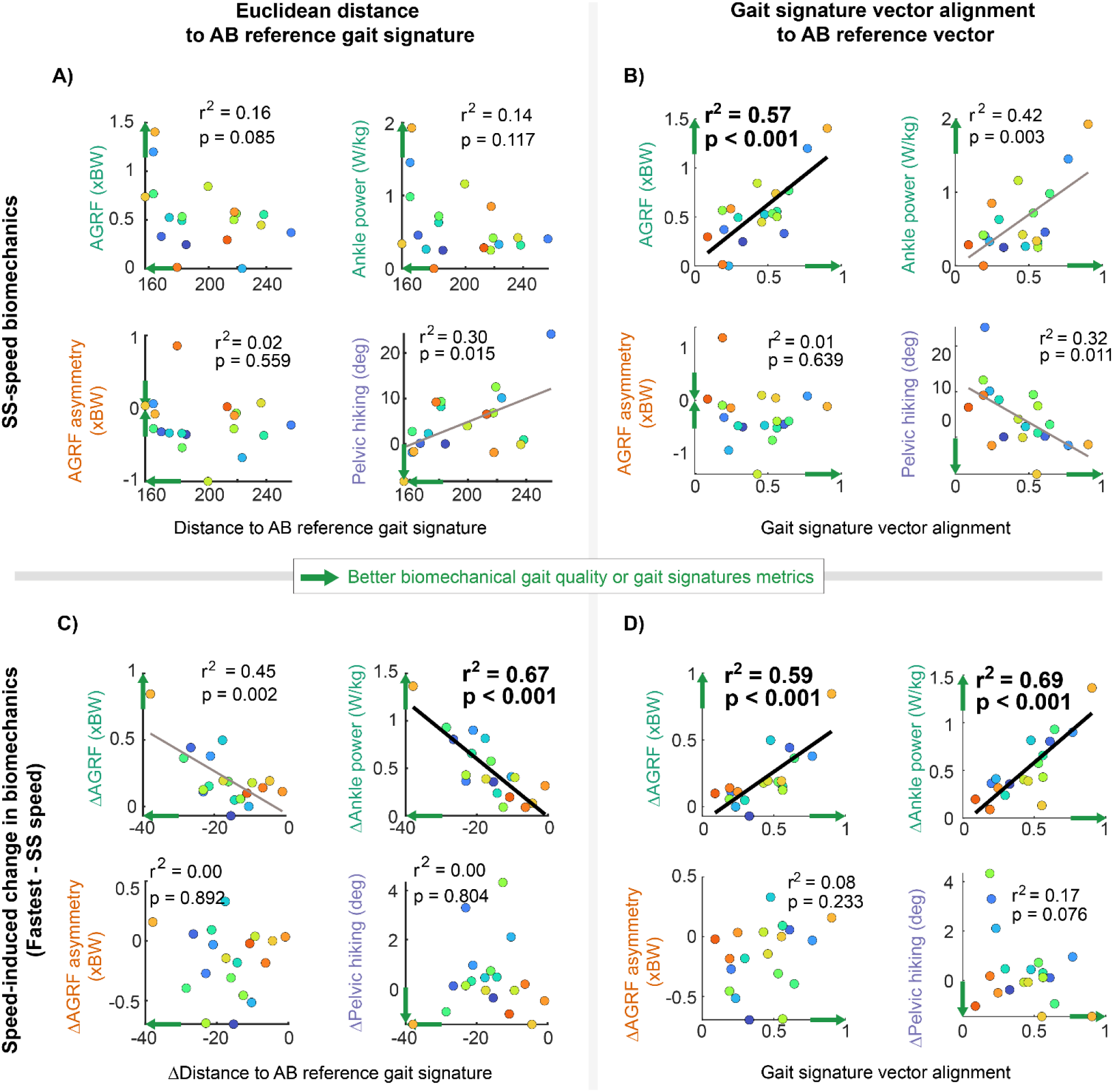
Correlations between post-stroke gait signatures metrics and discrete biomechanical variables. Colored dots denote post-stroke participants. The discrete biomechanical variables presented encompass significant correlations and examples of non-significant correlations in inter-limb asymmetry and compensation variables. Example correlations with discrete biomechanical variables are shown for the Euclidean distance to the AB reference gait signature (left column) and the alignment of gait signatures to the AB reference gait signature. Green arrows denote the direction of improvement in each biomechanical variable and the direction considered an improvement in the gait signatures metrics. Arrows are omitted from asymmetry variables, as the direction corresponding to improved (i.e., reduced) inter-limb asymmetry depends on SS-speed asymmetry. Thick black regression lines denote significant correlations after correction for multiple comparisons (α_Sidak_ = 9.8e-4). Thin black regression lines denote marginally significant correlations (p < 0.05). The upper panels assess the relationship between SS-speed discrete biomechanical variables and **A)** Euclidean distances between post-stroke gait signatures and the AB reference gait signature and **B)** post-stroke gait signature vector alignment to the AB reference vector. The lower panels assess the relationship between speed-induced changes (*Δ*: Fastest minus SS speed) in biomechanical variables and **C)** speed-induced changes in the Euclidean distances between post-stroke gait signatures and the AB reference gait signature and **D)** post-stroke gait signature vector alignment to the AB reference vector. Correlations with all discrete biomechanical variables can be found in *Supplemental Figures S4 & S5*. Abbreviations: AGRF = anterior ground reaction force

### Across walking speeds, gait signature similarity to an AB reference gait signature holistically encoded inter-individual differences in biomechanical gait quality

Because gait signatures and discrete biomechanical variables emerge from the same underlying neuromechanical properties, and because changes in different gait signatures dimensions (i.e., PCs) may capture different characteristic changes in gait biomechanics, we tested the ability of gait signatures PCs to predict multidimensional biomechanical variables across walking speeds (variables shown in Figure 6A, left). To test this hypothesis, we used Partial Least Squares (PLS) correlation analysis^20,28,29^, a methodology that identifies lower-dimensional subspaces that simultaneously describes two high-dimensional variables: the gait signature PCs and the discrete biomechanical variables.

**Figure 6:**
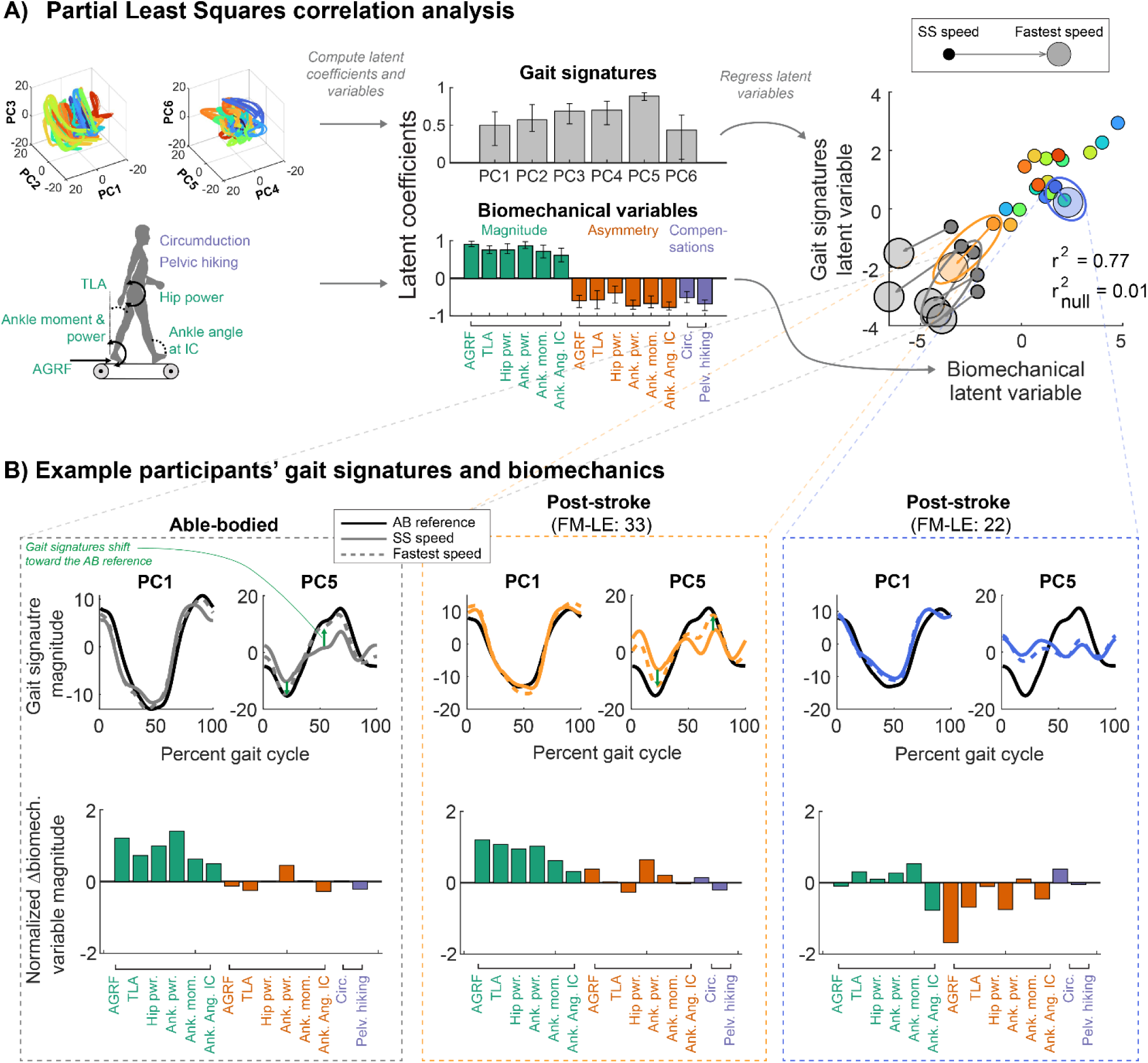
Holistic relationships between multiple gait signature PCs and multiple biomechanical variables across participants and speeds. **A)** Partial Least Squares (PLS) correlation analysis identified linear transformations of gait signatures PCs and biomechanical variables to maximize the covariance of the two datasets. Gait signatures variables consisted of the Euclidean distance between each participant’s first 6 PCs and those of the AB reference (PC traces shown, left). Biomechanical variables consisted of six paretic-leg magnitude variables (green), six asymmetry variables (orange), and two compensation variables (purple). For the latent coefficients (middle), the median and 95% confidence interval (1000 iterations of 4-fold cross validation) are shown for gait signatures (gray) and biomechanical variables (colors). The gait signatures and biomechanical latent variables (right) are shown for the SS speed (small dots) for all participants and the fastest speed (large dots) for AB participants (gray) and two example post-stroke participants (colors). For these participants, arrows connect the SS and fastest-speed dots. The three ellipses highlight the example participants shown in Panel B. The r^2^ value denotes the median out-of-sample correlation (4-fold cross validation) and r^2^_null_ denotes the median out-of-sample correlation, identified by applying PLS to randomly shuffled data. **B)** Example gait signatures and biomechanical variables for one AB participant (left) and two post-stroke participants (middle and right). Waveforms show stride-averaged gait signature PCs 1 & 5 at the SS (solid line) and fastest (dashed line) speed, and the AB reference gait signature (black). Green arrows highlight shifts toward the AB reference gait signature at faster speeds. Bar plots show the speed-induced change (fastest vs SS speed) in each of the 14 biomechanical variables. Bar colors match those described in Panel A. For clarity, each variable was normalized to unit variance across participants and speeds. Abbreviations: AGRF = anterior ground reaction force; Ank. Ang. IC = ankle angle at initial contact; Ank. mom. = peak ankle plantarflexor moment; Ank. pwr. = peak ankle plantarflexor power; Circ. = circumduction; FM-LE = lower-extremity Fugl-Meyer score; Hip pwr. = Peak hip power; PC = principal component; Pelv. Hiking = pelvic hiking; TLA = trailing limb angle.

Using gait signatures and biomechanical variables across walking speeds, the PLS model identified one significant latent dimension, consisting of latent coefficients (Figure 6A, middle) and the corresponding latent variables (Figure 6A, right; 4-fold cross-validation r^2^ = 0.77; p < 1e-16). The latent coefficients of this dimension reflected that more AB-like gait signatures corresponded to greater holistic biomechanical gait quality: The signs of the gait signatures coefficients (Figure 6A, middle, gray bars) relative to those of the biomechanical variables Figure 6A, middle, colored bars) reflected that gait signatures PCs that are more similar to the AB reference corresponded to increased paretic leg biomechanical output (green bars), decreased inter-limb asymmetry (orange bars), and decreased gait compensations (purple bars). The correlation between latent variables in this dimension are shown on the right of Figure 6A (latent variables at the SS speed are shown as small dots). Latent variables at the fastest speed (large dots; shown for AB adults and two post-stroke participants) show that the relationship between gait signatures and multiple biomechanical variables is consistent in AB adults (i.e., latent variables change in the same direction) but not in all stroke survivors. For example, one post-stroke participant with a higher lower-extremity sensorimotor function score (orange dots; FM-LE = 33) exhibited similar speed-induced changes in gait signatures and biomechanical latent variables to AB adults. Conversely, another post-stroke participant with a lower sensorimotor function score (blue dots; FM-LE = 22) exhibited speed-induced changes in gait signatures and biomechanical latent variables orthogonal to AB adults in this PLS latent dimension.

Figure 6B highlights how gait signatures (top) and biomechanical variables (bottom) differ at the SS and fast walking speeds for three example participants. For gait signatures, only PCs 1 & 5 are shown. PC 1 exhibited small changes with respect to speed, whereas PC 5 exhibited larger changes with speed (Figure 6B; solid vs. dashed lines). This speed-induced change is consistent with the latent coefficient corresponding to PC 5, which was larger and more stable than other coefficients (Figure 6A, middle). Between the SS (solid lines) and fastest speeds (dashed lines), gait signatures PCs 1 & 5 for one AB participant (left) and one post-stroke participant with a higher lower-extremity sensorimotor function score (middle; FM-LE = 33; orange ellipse in Figure 6A right) shifted toward the AB reference gait signature (black lines). Both the AB participant and this post-stroke participant exhibited improvements in right/paretic-leg biomechanical variables and relatively small changes in inter-limb asymmetry and gait compensations (Figure 6B; bar plots show change between the fastest and the SS speeds; variables are normalized for clarity). Conversely, speed-induced changes in gait signature PC 5 of a post-stroke participant with a lower sensorimotor function score (FM-LE = 22; Figure 6B, right; blue ellipse in Figure 6A right) were small. At faster speeds, this participant’s paretic-leg biomechanical magnitude variables improved only slightly, while inter-limb asymmetry worsened.

### Gait signatures predicted discrete gait biomechanical variables more holistically than walking speed alone, which did not capture individual differences in inter-limb asymmetry and gait compensations

The model that used walking speed to predict discrete biomechanical variables (speed-based PLS model) had similar latent coefficients in the first latent dimension as the gait signatures-based PLS model (Figure 7A; colored bars). The speed-based PLS model predicted individual differences in most paretic-leg biomechanical variables accurately (r^2^ = 0.15-0.87; Figure 7B, top left; green dots in Figure 7C, left column), but not inter-limb asymmetry and compensation variables (r^2^ = 0.01-0.21; Figure 7B, bottom left; orange and purple dots in Figure 7C, left column). Compared to the speed-based PLS model, the gait-signatures-based PLS model predicted paretic leg biomechanical variables slightly less accurately (r^2^ = 0.21-0.70) but predicted inter-limb asymmetry and compensation variables slightly more accurately (r^2^ = 0.13-0.32; Figure 7C, right column).

**Figure 7:**
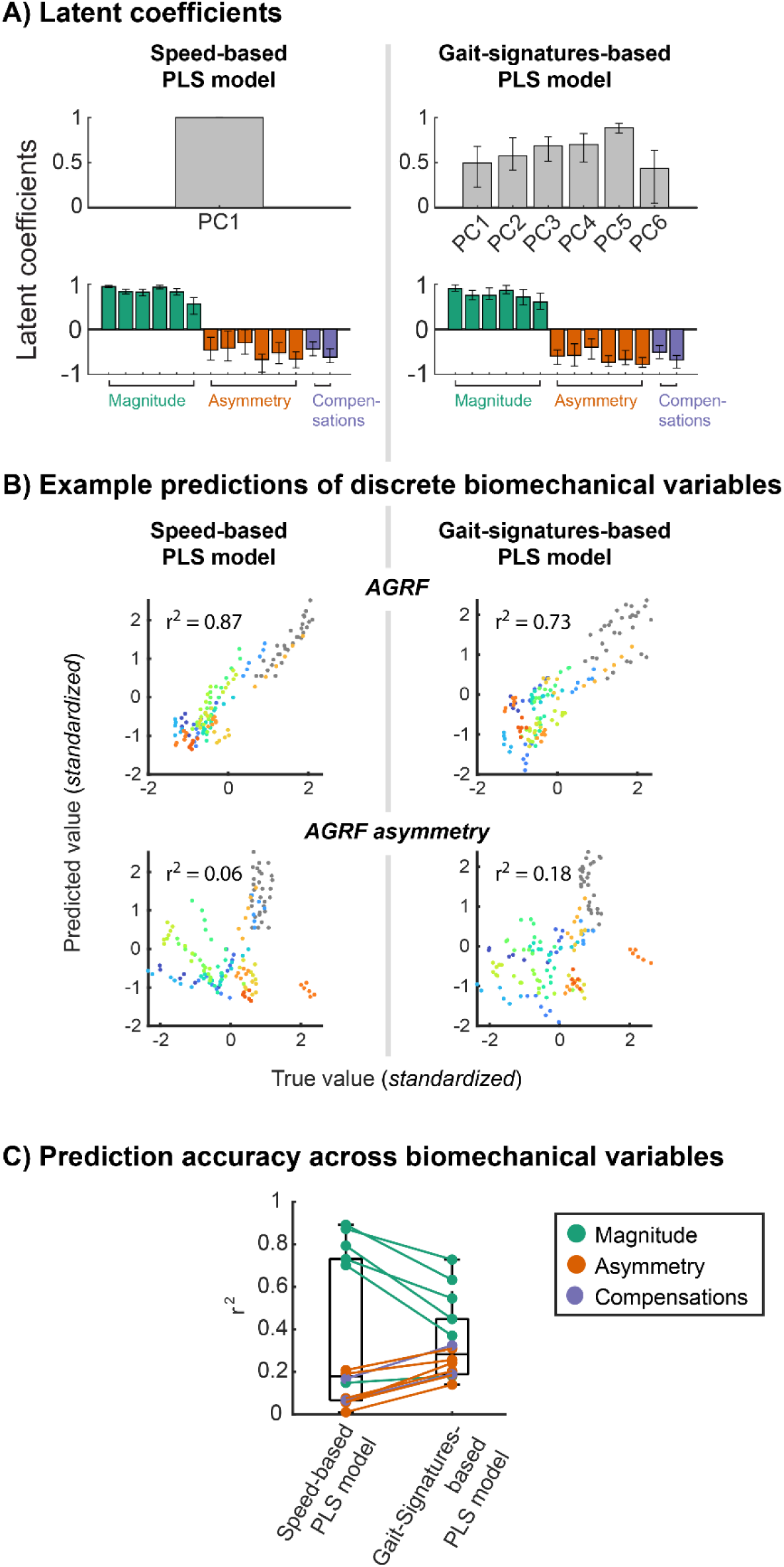
Simultaneous predictions of 14 discrete biomechanical variables from walking speed compared to predictions using gait signatures PCs. Predictions were generated using PLS models fit between the 14 discrete biomechanical variables and, separately, walking speed (“Speed-based PLS” model) and the first 6 gait signatures PCs (“Gait-Signatures-based PLS” model). Coefficients of determination (r^2^) for each variable represent the median model performance over 1000 iterations of 4-fold cross validation across all participants and walking speeds. A) PLS model latent coefficients for the speed-based (left) and gait-signatures-based (right) PLS models. Gray bars represent the variables that differ between the two models: the speed-based model contains only one latent coefficient corresponding to walking speed, while the gait-signatures-based model contains six coefficients corresponding to the gait signatures PCs. Colored bars represent the biomechanical variables, which were used in both models. **B)** Example predictions of paretic-leg AGRFs (top) and AGRF asymmetry (bottom). The left column shows speed-based predictions. The right column shows gait-signatures-based predictions using gait signatures PCs. Colors represent participants. **C**) Boxplot showing median coefficients of determination (r^2^) across cross validation iterations for each biomechanical variable for the Speed-based and Gait-Signatures-based PLS models. Green dots represent paretic-leg magnitude variables. Orange dots represent inter-limb asymmetry variables. Purple dots represent gait compensation variables. Lines connect dots representing the same variable in the two models. Abbreviations: AGRF = anterior ground reaction force. PLS = Partial Least Squares

## DISCUSSION

This study showed that neuromechanical gait signatures—latent representations of gait dynamics derived from timeseries joint kinematics using an RNN—holistically capture individual differences in post-stroke gait biomechanics across walking speeds. Post-stroke gait signatures becoming more like AB gait signatures at faster walking speeds suggests that faster speeds improve overall biomechanical gait quality. However, the *direction* that gait signatures change with speed (i.e., gait signature vector alignment) better reflects inter-individual differences in the neuromechanical dynamics governing post-stroke walking function and gait biomechanics than does the baseline similarity to AB gait signatures (i.e., SS-speed Euclidean distance)^20^. Our PLS correlation analysis demonstrates that the rich, multidimensional representation of gait provided by gait signatures enables a simple representation of holistic biomechanical gait quality, which may be useful in personalizing walking speeds during post-stroke gait retraining. While this work focused on walking speed, gait signatures may be extended to assess how overall gait quality changes with various treatments (e.g., functional electrical stimulation)^30,31^ or populations (e.g., people with cognitive impairment)^31–33^.

Our results suggest that post-stroke gait signatures capture how multiple aspects of gait biomechanics change with speed, such that gait signatures may be useful in quantifying biomechanical gait quality. Prior studies reported that walking at faster speeds typically improves biomechanical gait quality according to discrete biomechanical metrics including paretic-leg propulsion and joint mechanics, step length, and joint excursions^7–9,34^. Corroborating these findings, we interpret post-stroke gait signatures becoming more like AB gait signatures with increasing walking speed as suggesting that faster speeds improve biomechanical gait quality. Similar trends toward AB gait signatures at faster speeds in the paretic-leg stance and swing phases of the gait cycle support improvements in gait quality with speed over the entire gait cycle. This finding is consistent with the Gait Deviation Index (GDI), which is defined by the similarity of a low-dimensional representation of an individual’s stride-averaged kinematics to that of a corresponding AB reference^35^. In contrast with the GDI, encoding gait dynamics from continuous timeseries kinematics using the gait signatures RNN may capture spatiotemporal relationships spanning multiple timescales: from fractions of a second to multiple strides. We posit that these multi-timescale relationships may encode subtle—but important—aspects of biomechanical gait quality^36^. Further, gait signatures captured speed-induced improvements in biomechanical gait quality without requiring *a priori* selection of biomechanical variables to define gait quality and are not limited to measurements at specific timepoints^7–9,34,37^. Therefore, gait signatures may be useful across a range of contexts and research questions in which the measurements that best reflect gait quality are not known. However, our findings only support that post-stroke gait signatures encode *biomechanical* gait quality, as defined relative to an AB reference. Gait signatures may not encode other factors affecting gait quality or performance—such as the energy cost of walking or gait stability—which may not be improved by increasing the similarity of a stroke survivor’s gait signatures to those of AB adults due to the neuromechanical constraints underlying hemiparetic gait^24,38^. Characterizing the relationship between gait signatures and other aspects of gait quality and performance, or relative to other normative references, represents an exciting area of future research.

Our findings suggest that gait signature vector alignment, not the similarity of gait signatures to AB adults, encodes individual differences in the neuromechanical constraints shaping post-stroke gait biomechanics. This finding is consistent with prior studies that suggested that *how* post-stroke gait biomechanics change with speed reflects differences in neuromechanical deficits that are not revealed by SS-speed biomechanics^11,17^. The weak correlations between the similarity (i.e., Euclidean distance) of post-stroke gait signatures to AB gait signatures and walking speed may be due to aspects of neuromechanical dynamics that differ between individuals but do not constrain feasible walking speed, such as learned motor accents^39,40^. Further, our Euclidean distance metric does not reflect *how* gait signatures differ between individuals, which may be too coarse to capture relationships between gait signatures and gait biomechanics. In contrast, gait signature vector alignment quantifies the similarity of *how* post-stroke gait signatures change with speed relative to AB gait signatures. Therefore, gait signature vector alignment may better encode the neuromechanical constraints (e.g., altered muscle coordination^41^) or movement goals (e.g., energy cost minimization^38^) that underlie how stroke survivors modulate gait biomechanics across speeds. Alternatively, gait signature vector alignment may reflect the mechanical constraints of walking faster: it may be difficult to walk faster while changing gait signatures differently from AB adults^21^. Such mechanical constraints may explain correlations with *speed-induced changes* in individual paretic-leg biomechanical variables, but not correlations with biomechanical variables at participants’ SS speeds^16,23,42,43^. Gait signature vector alignment does not capture constraints that shape individual differences in inter-limb asymmetry^11,16^. Therefore, a limitation of the walking speed modulation paradigm is that it is insufficient to disentangle the neuromechanical and mechanical factors captured by gait signatures. For example, walking faster by increasing, maintaining, or decreasing inter-limb asymmetry may have differential effects on gait signatures.

Alternative experimental paradigms that modulate mechanical constraints are needed to overcome this limitation.

The rich, multidimensional representation of gait provided by gait signatures captures holistic biomechanical gait quality and may be useful in personalizing walking speeds during post-stroke gait retraining. To identify the impacts of speed on post-stroke gait biomechanics, prior studies often used linear regression or statistical hypothesis tests (e.g., t-tests) across multiple biomechanical variables^7–9,44,45^. Consistent with these studies, we found that, compared to individuals’ SS speeds, their fastest safe walking speed improved paretic-leg biomechanical variables and had individual-specific impacts on inter-limb asymmetry and gait compensations. Gait signatures appear to balance these effects: gait signatures were most similar to AB gait signatures at the fastest speed for only 58% of participants, between the percentages for individual paretic leg biomechanical variables (≥58%) and inter-limb asymmetry and compensation variables (≤26%). More recent machine learning methods (e.g., k-means clustering; convolutional neural networks) identified non-trivial effects of walking speed on inter-limb asymmetry within sub-groups of stroke survivors despite variable changes in gait kinematics or kinetics^16,42,46^. Our gait-signatures-based PLS model results corroborate this work by identifying a latent dimension that reflects improved gait quality across paretic-leg biomechanics, inter-limb asymmetry, and gait compensation variables.

Our gait-signature-based PLS modeling approach may be useful for selecting gait speeds during training due to its interpretable and continuous representation of biomechanical gait quality. For example, the PLS model’s latent coefficients revealed the relative importance of each gait signature PC on gait biomechanics. These coefficients suggested that gait signature PC 5, not PC 1, may serve as an indicator of how walking speed modulation impacts holistic biomechanical gait quality. Our prior findings suggested that PC 1 captures basic limb alternation during walking that span individuals^20^. In contrast, PC 5 represents a smaller amount of variance in the gait signatures and may encode neuromechanical idiosyncrasies that shape individual differences in gait biomechanics. Further, selecting walking speeds based on the rich representation of neuromechanical dynamics provided by gait signatures may lead to better overall biomechanical gait quality than would be achieved by maximizing walking speed^16,23,42,43^. While increasing walking speed can be used to predict individual differences in paretic-leg biomechanics, the additional information contained in gait signatures PCs is needed to predict changes in both paretic-leg inter-limb asymmetry and gait compensations. A limitation of our PLS approach is that we identified only one latent dimension, which cannot identify responder sub-groups that may exhibit characteristic changes in gait signatures and biomechanics with speed^46^.Training a gait-signatures-based PLS model on larger sample sizes may identify additional latent dimensions reflecting characteristic responses to walking speed^41,42,46^.

While similarly interpretable models exist, to our knowledge they have not been used to analyze gait speed modulation in stroke survivors^47^. The gait-signature-based PLS latent variables extend beyond clustering and discrete group labels, acknowledging the continuous nature of post-stroke impairments and may be useful in informing therapy parameters for individuals on the margins of an impairment sub-group^42,46^. Our latent states are analogous to those of a recent convolutional neural network model, which could similarly inform therapy parameters^46^. However, this convolutional framework identified latent states to best classify stroke survivors versus AB adults at preferred walking speeds, rather than to encode neuromechanical gait dynamics across multiple walking speeds. Alternative metrics of gait coordination^47,48^ and quality^35,49^ exist and could be compared against gait signatures to determine which features and frameworks best inform gait speed selection.

Our findings provide preliminary support for gait signatures as a potential biomarker to assess how overall gait quality changes across treatments and populations. By encoding neuromechanical gait dynamics from sagittal-plane joint kinematics, the gait signatures RNN is designed to be a generalizable representation of gait that may be useful in various contexts, without requiring force plates or even 3D kinematics to capture individual differences in both gait neuromechanics^20^ and 3D gait biomechanics. One limitation of using sagittal-plane kinematics is that 3D kinematics may better encode frontal-plane gait deviations (e.g., gait compensations) in stroke survivors, potentially strengthening relationships between gait signatures and biomechanical variables^7,16,21,45^. However, requiring only sagittal-plane kinematics supports its feasibility for video-based gait analysis in lower-resource settings^50^. Further, our findings are limited to the context of post-stroke gait speed modulation. Future work should validate the utility of gait signatures for other therapeutic paradigms and patient populations. For example, gait signatures may be useful in quantifying the effects of other therapies, such as functional electrical stimulation or dance-based movement therapy, on movement quality^30–32^.

Gait signatures may also be useful in holistic assessment of movement quality in other populations, including people with Parkinson’s disease or cognitive impairment^32,51^ or in more complex movements like those performed by athletes^33^. Collectively, our findings support further development of gait signatures in other application areas and populations, and using larger sample sizes to understand how neuromechanical constraints shape gait signatures.

## METHODS

Twenty individuals post-stroke (6 female; 61 ± 10 years; 44 ± 49 months post-stroke; Table 1) and 5 able-bodied adults (4 female; 24 ± 4 years; Table 1) participated in one session of treadmill-based gait analysis. Study procedures were approved by the Emory University Institutional Review Board and all participants provided written informed consent. Inclusion criteria included >6 months post-stroke (not applicable for AB participants), the ability to walk on a treadmill without an orthotic device for 1-minute, and the ability to communicate with investigators. Exclusion criteria included neurologic diagnosis other than stroke, hemi-neglect, orthopedic conditions limiting walking, and cerebellar dysfunction. For post-stroke participants, a trained clinician evaluated overground self-selected (SS) walking speed using the 10-meter walk test and lower-limb sensorimotor impairment using the lower-extremity Fugl-Meyer (FM-LE; Table 1) before treadmill-based gait analysis^52,53^. Study procedures were approved by and performed in accordance with guidelines set forth by the Emory University Institutional Review Board. All participants provided written informed consent.

**Table 1:**
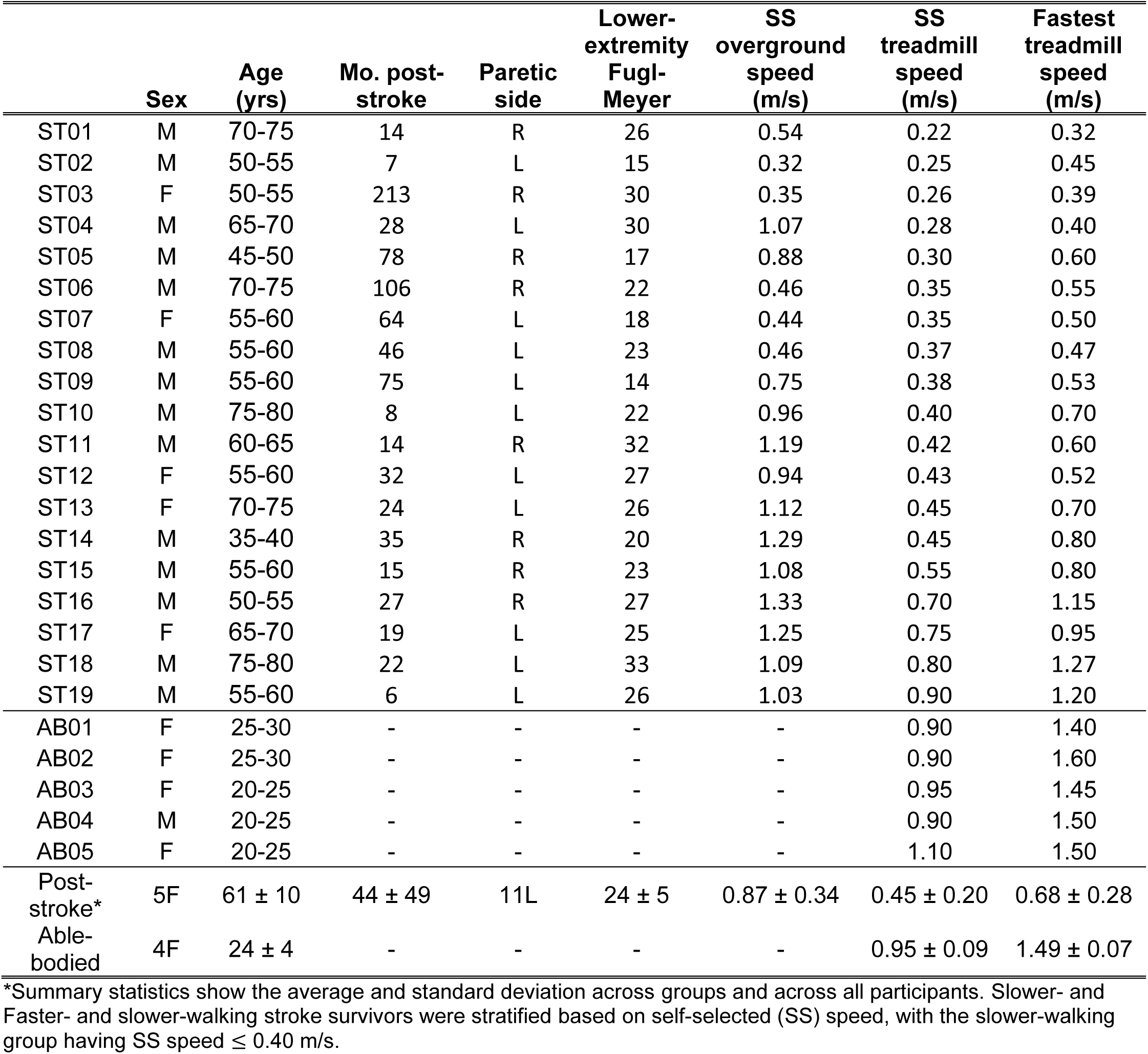
Participant demographics.

### Experimental setup

Participants walked on a split-belt instrumented treadmill (Bertec Corp., Ohio, USA) to enable collection of ground reaction force (GRF) data independently for each limb at 1000 Hz. Reflective markers were attached to the trunk, pelvis, and bilateral thigh, shank, and foot segments^37^. Marker trajectories were recorded at 100 Hz using a 7-camera motion capture system (Vicon Inc., Oxford, UK). During all walking trials, for safety, participants held onto a front handrail and wore an overhead safety harness without body-weight support. Participants were instructed to maintain a light and consistent handrail grip for all walking trials; if investigators suspected a change in handrail grip or excessive reliance on the handrail, the participant was given feedback and, if needed, the trial was restarted.

### Determination of walking speeds

After a 30-60 second trial to acclimatize to the treadmill, each participant’s self-selected (SS) walking speed was determined by slowly increasing the belt speed until the participant reported their comfortable walking speed. Next, the fastest safe walking speed was determined by gradually increasing the treadmill speed above their SS speed until either the participant reported the fastest speed that they could safely walk for 30 seconds, or the investigators deemed it unsafe to increase the speed further. Next, four intermediate speeds were computed at equal increments between the SS and fastest walking speeds, resulting in a total of six evenly distributed speeds spanning each participant’s walking capacity. Data were collected during 15-second treadmill walking trials at each of the six speeds, in increasing order from the SS speed to the fastest speed. Brief 1–2-minute standing rest breaks were provided between walking trials as needed, to prevent fatigue. Based on SS speed, participants were stratified into slower-(SS speed ≤ 0.4 m/s) and faster-walking (SS speed > 0.4 m/s) groups^8,9,27^.

### Data processing

All GRFs, joint kinematics, and joint moments and powers were processed in Visual 3D (C-Motion Inc., Maryland, USA). Before computing joint moments and powers, GRFs and joint kinematics were low-pass filtered at 30 and 6 Hz, respectively using a 4^th^-order Butterworth filter. At each gait speed, dependent variable magnitudes were computed for the paretic and non-paretic limbs. For each variable, the average value across all gait cycles during each trial was used for each speed.

### Discrete biomechanical variables

To holistically characterize post-stroke gait biomechanics across walking speeds, we computed 14 discrete biomechanical variables including the magnitude of 6 paretic-leg kinematic and kinetic variables, 6 inter-limb asymmetries variables, and 2 gait compensation variables using Visual 3D (C-Motion, USA). Paretic leg magnitude variables included: Peak paretic propulsion was normalized to body mass and was calculated as the peak value of the anteriorly-directed GRF during the terminal double-support phase of the limb^54,55^. Trailing limb angle (TLA) was calculated as the maximum angle between the vertical axis of the laboratory and a line joining the greater trochanter and fifth metatarsal head marker^56–58^. Peak ankle moment and peak ankle power were calculated during the stance phase of gait. Peak hip power was defined as the peak hip flexor power during the pre-swing and initial swing phase of gait. Peak moments and powers were normalized to body mass^8,54^. Additionally, ankle angle at initial contact (IC) was calculated. Inter-limb asymmetry variables included the inter-limb asymmetry of each paretic-leg magnitude variable, calculated as the difference between the non-paretic and paretic legs^59^. We included two gait compensation variables: hip circumduction and pelvic hiking of the paretic leg. Paretic leg circumduction was calculated as the maximum frontal plane deviation of the bottom heel marker during stance phase versus the subsequent swing phase. Pelvic hiking was calculated as the maximum frontal plane angle between the pelvis during a static standing calibration trial and during the swing phase of the paretic leg^7^. The dataset containing participant demographics and discrete biomechanical variables used in this study can be found in https://github.com/bermanlabemory/GaitSignatures_PostStrokeGaitBiomechanics.

### Gait signatures

#### Gait signatures recurrent neural network training

To model gait signatures as a dynamic representation of the neural and biomechanical constraints governing movement, we trained a single recurrent neural network (RNN) to predict the time-evolution of sagittal-plane leg joint kinematics for all participants and walking speeds. That is, the RNN predicted a one-time-step shifted version of bilateral sagittal-plane hip, knee, and ankle kinematics. As in our previous work, the network consisted of one long short-term memory (LSTM) hidden layer with 512 nodes and a lookback window of 499 samples^20^. During model training, the data were split into training (67% of the data per trial) and validation (33% of the data per trial) sets. The RNN was trained using Keras 2.15.0 and Tensorflow 2.15.0. Model training used the ADAM optimizer with a learning rate of 0.0001. Training was conducted on batches of data corresponding to individual trials, with a loss function minimizing the mean squared error of single-timestep predictions. Convergence was determined after 10000 epochs or when the validation error did not decrease in 200 iterations. All 6 speeds for 24 participants were used to train the RNN. The trials were concatenated along the time dimension. To capture gait signatures associated with the paretic-leg, we used the convention that the paretic legs of post-stroke participants were equivalent to the right legs of the AB participants. The RNN can discover participant-and trial-specific representations of walking because the LSTM’s latent activation and cell states (*H_t_* and *C_t_*, respectively, in Figure 1) are updated from participant-and trial-specific timeseries data. These latent states exist on the manifold over which each participant’s gait evolved.

To compute participant-and trial-specific gait signatures, we extracted the time-varying latent states of the network using the entire timeseries for each trial (1500 samples/trial). Because each LSTM node contains one activation and one cell state, each trial’s latent states had dimensions [1024 x time]. We computed the gait signatures by applying Principal Components Analysis (PCA) to the latent states for all trials, concatenated along the time axis. The first 6 PCs were used to define the gait signatures, accounting for 92% of the variance in the latent states across trials (*Supplemental Figure S1*). Finally, to produce stride-averaged gait signatures, we phase-averaged the gait signatures by first estimating the continuous phase of each trial using Phaser^26^. We phase-averaged the gait signatures using a weighted sum of all samples in the trial, weighted according to a von Mises distribution based on their proximity to each target phase value over the gait cycle^60^. To ensure consistent comparisons of gait signatures across trials, phase estimates were aligned for each participant by first setting phase of the participant’s SS-speed trial to 0 radians at initial contact of the right (AB participants) or paretic (post-stroke participants) leg. To ensure alignment across trials, we then aligned all trials for each participant to their SS-speed trial by minimizing the difference between PC 1 of the SS-speed gait signature and PC 1 of all other gait signatures. These phase-averaged PCs constituted the gait signatures and were used to compute the metrics described below. Due to data corruption on one trial, one participant, ST16, only had five walking speed trials available, with the fastest walking speed missing. This participant’s data were used when training the gait signatures RNN, but were not included in subsequent analysis.

#### Gait signatures metrics

To compare gait signatures to AB reference gait signatures, we separately computed the average AB gait signatures at each walking speed condition: from the SS speed to the fastest speed (smaller black squares in Figure 1D). We found that the average AB gait signature changed linearly with speed (median r^2^ = 0.82, computed across the first 6 gait signatures PCs)^21^. To ensure that AB participants also moved toward the AB reference with increasing walking speed, we defined the AB *reference* gait signature as the extrapolation of the average AB adult gait signatures to 1.75 m/s, 17% faster than the average fastest AB walking speed (1.50 m/s; *Supplemental Figure S2).* The extrapolated AB reference gait signature is represented by a large black square on the gait maps in Figure 1D.

To capture the relative similarity of gait signatures we computed the Euclidean distance between each pair of trials, the AB reference gait signature, and the AB reference gait signature at each speed, producing a dissimilarity matrix with dimensions 151×151. We constructed the gait maps in Figure 1 by applying 2-dimensional multidimensional scaling to the dissimilarity matrix^20^.

To capture how similar the direction of speed-induced changes in gait signatures were to AB adults, termed *gait signature vector alignment*, we first defined an AB reference gait signature unit vector. The AB reference vector was defined as a unit vector between the average AB gait signatures and the SS and fastest speeds. We then computed an analogous unit vector for each participant’s gait signatures. Gait signature vector alignment was defined as the dot product of each participant’s vector and the AB reference vector. Consequently, participants whose gait signatures changed with speed in a direction parallel or orthogonal to that of the average AB adult gait signatures would have gait signature vector alignment scores of unity or zero, respectively.

### Statistical analyses

Unless otherwise noted, we used two-tailed parametric statistical tests for all tests of group differences when the data did not violate assumptions of normality and homogeneity of variances. All statistical analyses were conducted using MATLAB 2021b (Mathworks Ltd, Natick, USA).

#### Characterizing the group and individual-level differences in gait signatures

To determine how post-stroke gait signatures differ from those of AB adults, we first compared the Euclidean distance of gait signatures to the AB reference gait signatures between post-stroke participants and AB participants using Mann-Whitney U tests (α = 0.05). To confirm that gait signatures were individual-specific, we compared the within-versus between-participant variability of gait signatures using Mann-Whitney U tests (α = 0.05). We computed within-participant variability as the average Euclidean distance between all pairs of gait signatures across walking speeds. We computed between-participant variability as the average Euclidean distances between one participant’s gait signatures and all other participants’ gait signatures, at each relative walking speed. To determine how speed-induced changes in post-stroke gait signatures differ from those of AB adults, we compared gait signature vector alignment between post-stroke participants and AB participants using Mann-Whitney U tests (α = 0.05).

#### Identifying relationships between post-stroke gait signatures and clinical measures of walking function and sensorimotor impairment

To identify relationships between the post-stroke gait signatures and clinical measures of walking function and sensorimotor impairment, we performed independent linear regressions between our two gait signatures metrics (*Euclidean distance to AB reference* and *vector alignment*) and the following clinical outcomes for post-stroke participants: (1) treadmill-based SS walking speed, (2) overground SS walking speed, (3) range of treadmill walking speeds (fastest speed – SS speed), and (4) lower-extremity Fugl-Meyer scores (FM-LE; max = 34). We report coefficients of determination (r^2^) and p-values of the regression slopes (Wald tests). Regression fits were considered significant only after correction for multiple comparisons (Holm-Sidak stepdown correction; initial α = 0.05)^61^. For all statistical analyses involving multiple comparisons, we report α_Sidak_ as the significance level beyond which no additional comparisons were significant. Non-significant comparisons with p < 0.05 are described as “marginally significant.”

#### Characterizing the impacts of walking speed on gait signatures

To characterize how increasing walking speed impacted post-stroke gait signatures, we first compared the similarity of post-stroke gait signatures to the AB reference gait signature between the SS and fastest walking speeds using Wilcoxon signed-rank tests (α = 0.05). We then determined the walking speed that minimized each participant’s gait signature Euclidean distance to the AB reference gait signature. We computed the percentage of participants for whom gait signature Euclidean distances were minimized at each relative walking speed. To qualitatively assess how absolute walking speed differentially affected gait signatures in slower-and faster-walking stroke survivors, we visualized the relationship between absolute walking speed and gait signature distances to the AB reference gait signature in each gait phase. To determine if walking speed had differential effects on post-stroke gait signatures in different phases of the gait cycle, we performed the above analyses over the full gait cycle, only the stance phase of the paretic leg (*stance-only*), and only the swing phase of the paretic leg (*swing-only*). Gait signatures with lower Euclidean distances to the AB reference gait signature were considered to have higher quality^35^.

#### Characterizing the impacts of walking speed on biomechanical gait quality

To characterize how increasing walking speed impacted post-stroke biomechanical gait quality, we determined the walking speed that maximized the quality of each of the 14 discrete biomechanical variables for each post-stroke participant (yellow dots in Figure 4). For each variable, we computed the percentage of participants for whom each variable’s quality was maximized at each speed. For paretic-leg biomechanical magnitude variables, increasing the magnitude of each variable with speed corresponded to increased biomechanical gait quality (e.g., increasing paretic-leg AGRFs)^8,18,19^. For inter-limb asymmetry variables, decreasing the absolute value of each variable with speed corresponded to increased biomechanical gait quality^7,9,11^. For compensatory variables, decreasing compensations with speed corresponded to increased biomechanical gait quality^7,9^.

#### Identifying relationships between gait signatures and discrete biomechanical variables

To determine how post-stroke gait signatures were related to gait biomechanics at baseline (i.e., SS-speed) and changes in biomechanics with speed (i.e., Fastest vs SS speed), we performed univariate linear regression between our two gait signatures metrics (*Euclidean distance to AB reference* and *vector alignment*) and each biomechanical variable, separately. We report coefficients of determination (r^2^) and p-values of the regression slopes (Wald tests). Regression fits were considered significant only after correction for multiple comparisons (Holm-Sidak stepdown correction; initial α = 0.05)^61^.

#### Identifying holistic relationships between gait signatures and discrete biomechanical variables across walking speeds

To determine if gait signatures encoded a holistic (i.e., multidimensional) representation of post-stroke biomechanical gait quality across walking speeds, we used Partial Least Squares (PLS) regression to identify relationships between the first 6 PCs of gait signatures and all 14 discrete biomechanical variables. PLS identifies separate transformations of basis (i.e., *latent coefficients*) for two datasets in which the transformed variables (i.e., *latent variables*; see Figure 6A for a depiction of latent coefficients and variables) maximally covary^28,29^. The latent coefficients represent the relative weight of each feature on the corresponding latent variables. PLS produces multiple latent dimensions that, if generalizable and stable, would represent different relationships between gait signatures and discrete biomechanical variables^29^. We selected PLS regression because it performs well with small sample sizes and colinear variables^29,62^.

To enable identification of inter-and intra-individual relationships between gait signatures and biomechanics, we included data from all 6 walking speeds and all post-stroke and AB participants. For the gait signatures features, we used the Euclidean distance of each of the first 6 gait signatures PCs to the corresponding PCs of the AB reference gait signature. For discrete biomechanical variable features, we used the same variables described previously. All variables were z-scored before fitting the PLS model. All PLS steps were conducted using the *rCCA* function (regularization = 1.0) within the *cca-zoo* 2.6.0 library in Python 3.10.12^63^.

We analyzed PLS latent coefficients, latent variables, and correlations between the latent variables only for latent dimensions that were generalizable and stable. We considered a latent dimension generalizable if the out-of-sample correlation (r^2^) from 1000 iterations of 4-fold cross-validation was significantly greater than that of the first latent dimension of a null model (one-sided Mann-Whitney U tests; 1000 iterations with 30 samples per iteration; α = 0.05)^28^. The null model was fit using the same PLS procedure, but with the gait signatures and biomechanical variable samples randomly shuffled, without replacement, for each cross-validation iteration^62^. We considered a dimension stable if the 95% confidence interval of the latent coefficients over 1000 iterations of bootstrapping the model fitting procedure did not span the zero line^29^. Because the sign of PLS coefficients is arbitrary, we determined the sign of the coefficients of each cross-validation iteration that best matched those of the first iteration using cosine similarity, before computing the median and 95% confidence interval. We report the median and 95% confidence interval of latent coefficients, the median latent variables, and median out-of-sample correlations for the first latent dimension.

#### Determining if gait signatures holistically capture gait biomechanics

To determine if PLS regression using gait signatures captured holistic individual differences in gait biomechanics more accurately than walking speed alone, we compared the ability of a PLS model fit between discrete biomechanical variables and walking speed (speed-based model) to the model described above (gait-signatures-based model) to predict discrete biomechanical variables across walking speeds.

The speed-based and gait-signatures-based PLS models used the same model fitting procedure. PLS identified a single significant latent dimension with latent coefficients 𝐴 ∈ ℝ^𝑛×1^ and 𝐵 ∈ ℝ^𝑝×1^, and latent variables 𝑈 ∈ ℝ^𝑚×1^ and 𝑉 ∈ ℝ^𝑚×1^. For both models, *A* and *U* correspond to either the speed features (walking speed) or gait signatures features, while *B* and *V* correspond to the discrete biomechanical variable features. Here, *m* = 144 represents the number of samples in the dataset. The variable *n* represents the number of input features: *n* = 1 for the speed-based model and n = 6 for the gait-signatures-based model. The variable *p* = 14 represents the number of discrete biomechanical variable features. Here, the relationship between the data and the latent variables was represented as:

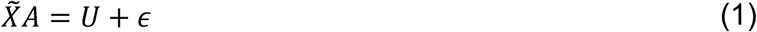

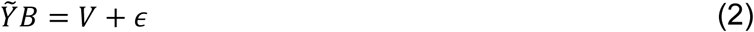

Where 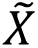 ∈ ℝ^𝑚×𝑛^ is a rank-one approximation of the speed or gait signatures features, 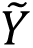 ∈ ℝ^𝑚×𝑝^ is a rank-one approximation of the discrete biomechanical variable features, and 𝜖 represents model error. To generate predictions, we fit a linear model in this one-dimensional latent space:

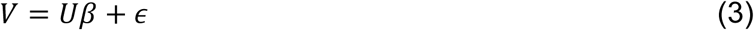

Where 𝛽 ∈ ℝ^1×1^ is a scalar coefficient. Next, we predicted the biomechanical latent variables, 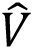 ∈ ℝ^𝑚×1^, based on the gait signatures latent variable, 𝑈 ∈ ℝ^𝑚×1^.

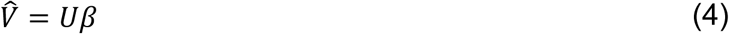

We generated predictions of the biomechanical variables, 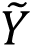 ∈ ℝ^𝑚×𝑝^, by projecting the predicted latent variables into the original basis of the discrete biomechanical variables:

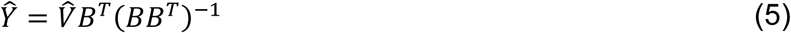

To evaluate model accuracy, we compared the coefficients of determination (*r^2^*) between the predicted and ground true discrete biomechanical variables. The reported r^2^ values represent the median r^2^ over 1000 iterations of 4-fold cross validation. We then compared model accuracy across paretic-leg magnitude variables and, separately, across interlimb asymmetry and compensation variables using signed-rank tests (α = 0.05).

## DECLARATIONS

### Consent for publication

Not applicable.

### Data availability

The datasets generated during and/or analyzed during the current study are available in the https://github.com/bermanlabemory/GaitSignatures_PostStrokeGaitBiomechanics.

### Authors’ contributions

M.R. drafted the original manuscript. M.R., G.B., L.T., and T.K. designed analyses. M.R. performed analyses and created all figures. T.K. collected the data. M.R., T.W., and T.K. processed the data and performed preliminary analyses that were foundational to the manuscript. M.R., L.T., and T.K. interpreted the data. T.K. was involved with all aspects of the study, including design, data collection, data processing, analysis, interpretation, and manuscript preparation. All co-authors contributed to critical review and revisions of the manuscript.

## Data Availability

All data produced in the present study are available upon reasonable request to the authors

## Acknowledgements

The authors would like to acknowledge Hanna Christianson, Justin Liu, Vincent Santucci, Payton Sims, Alex Schilder, and Laura Zajac-Cox for support in collecting the data used in this manuscript.

