## Supplemental for "Neuromechanical gait signatures reveal holistic biomechanical responses to walking speed modulation in stroke survivors"

### Supplemental – S1: Dynamical gait signatures capture holistic changes in post-stroke gait biomechanics with walking speed

**Authors:**

Michael C. Rosenberg[1*], Taniel S. Winner[1], Gordon J Berman[2], Lena H. Ting[1,4], Trisha M. Kesar[3,4]

[1]Department of Biomedical Engineering, Emory University & Georgia Institute of Technology, Atlanta, GA, USA

[2]Department of Biology, Emory University, Atlanta, GA, USA

[3]Department of Rehabilitation Medicine, Emory University School of Medicine, Atlanta, GA, USA

[4]Center for Physical Therapy and Movement Science, Emory University, Atlanta, GA

*Corresponding author

**KEYWORDS**

Biomechanics, gait rehabilitation, walking speed, gait dynamics, stroke, treadmill training

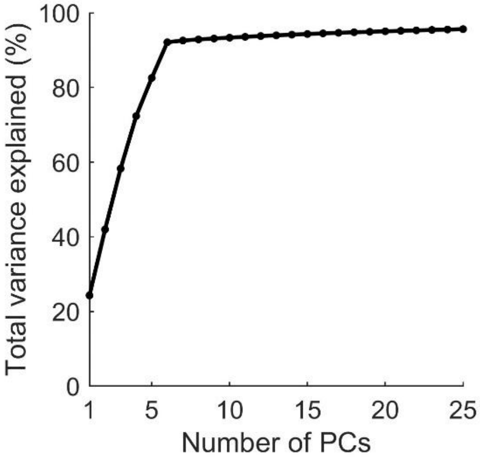


**Figure S1: Scree plot of the total variance explained by the first 25 gait signatures principal components (PCs)**. The N$^{th}$ sample represents the total variance explained by PCs 1-25.

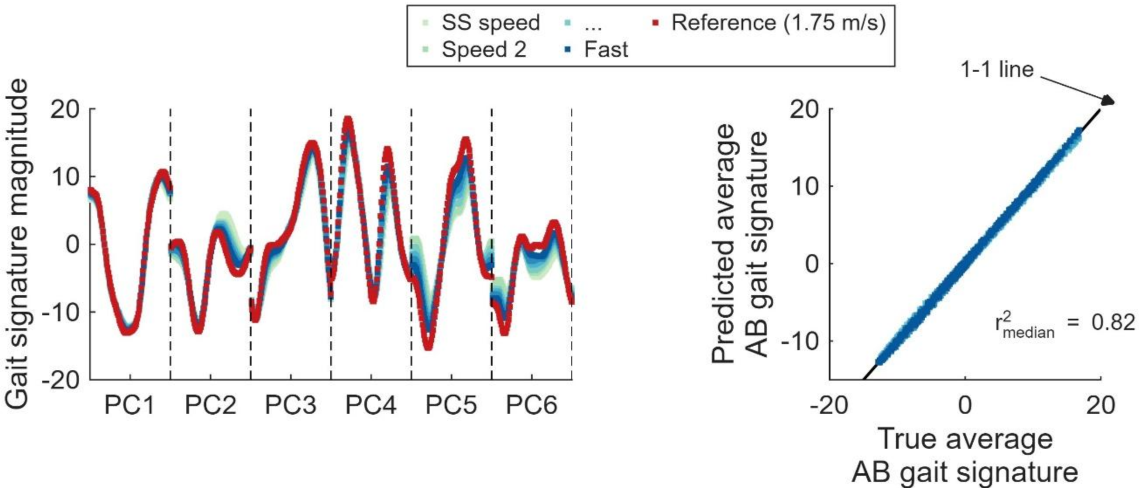


**Figure S2: Linear fit of the average able-bodied adult participants' gait signatures across the six walking speeds**. Left: The average AB gait signatures for the first 6 principal components (PCs) at each speed (blue hues) and the AB reference gait signature. Each PC contains 100 samples per stride. The AB reference gait signatures is a linear extrapolation of the gait signatures to 1.75 m/s. Right: Comparison of the true average AB gait signature at each of the 6 speeds (blue hues) and the average singature predicted by a linear fit between walking the average speed and the average gait signature. The solid black line denotes a perfect fit. Samples correspond to each sample of the first 6 gait signatures PCs (600 samples per speed). The $r^2$ value denotes the median coefficient of determination of speed predicting gait signatures across each of the 600 samples of the gait signature (100 samples per PC × 6 PCs).

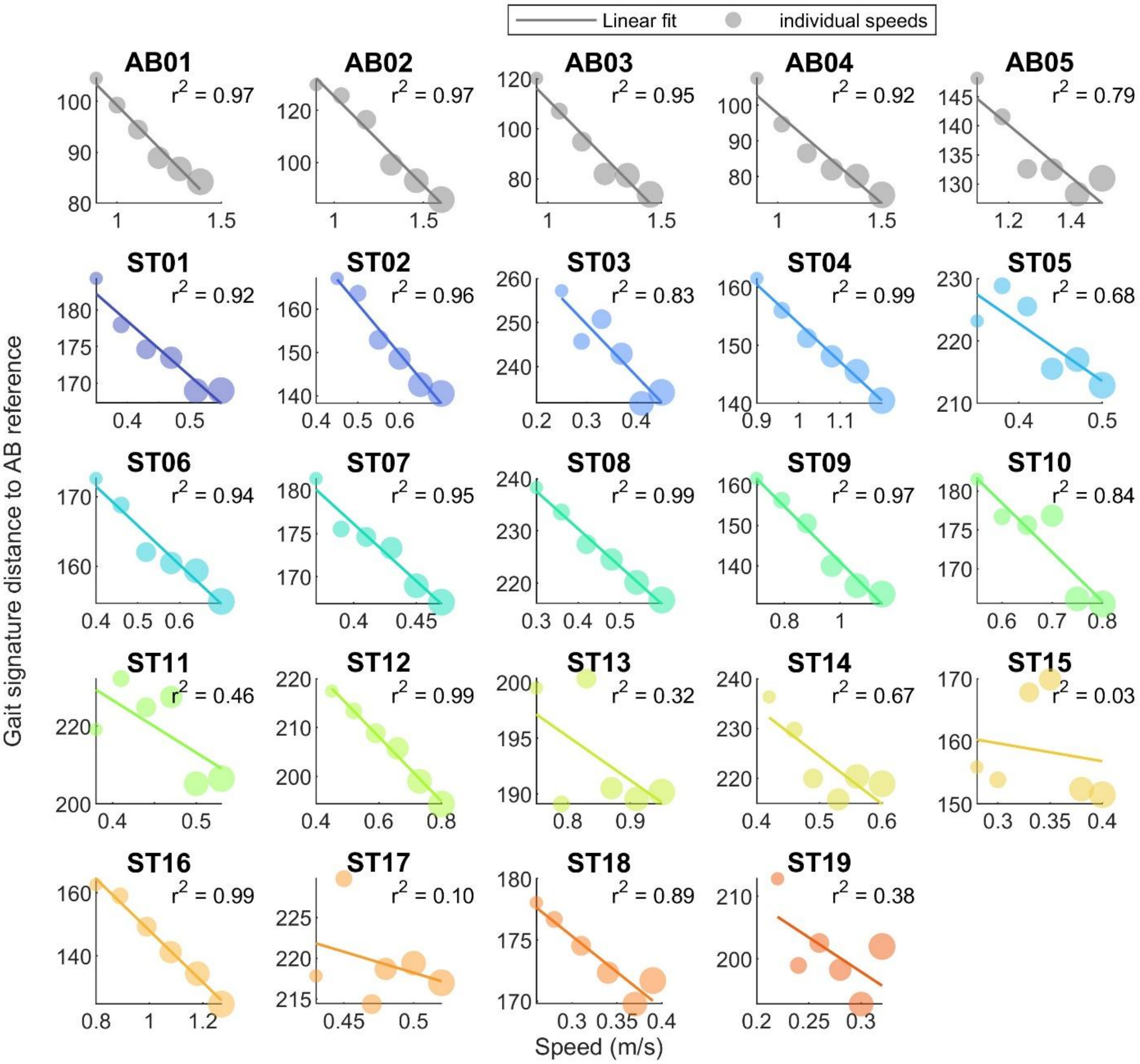


**Figure S3: Participant-specific changes in gait signature distances to the AB reference gait signature with walking speed**. Dots denote gait signatures corresponding to each walking speed. Larger dots denote trials at faster walking speeds. Sollid lines represent a linear fit between walking speed and each participant's gait signature. Not that the axis range varies for clarity.

**Gait signature metrics regressed against SS-speed discrete biomechanical variables.**

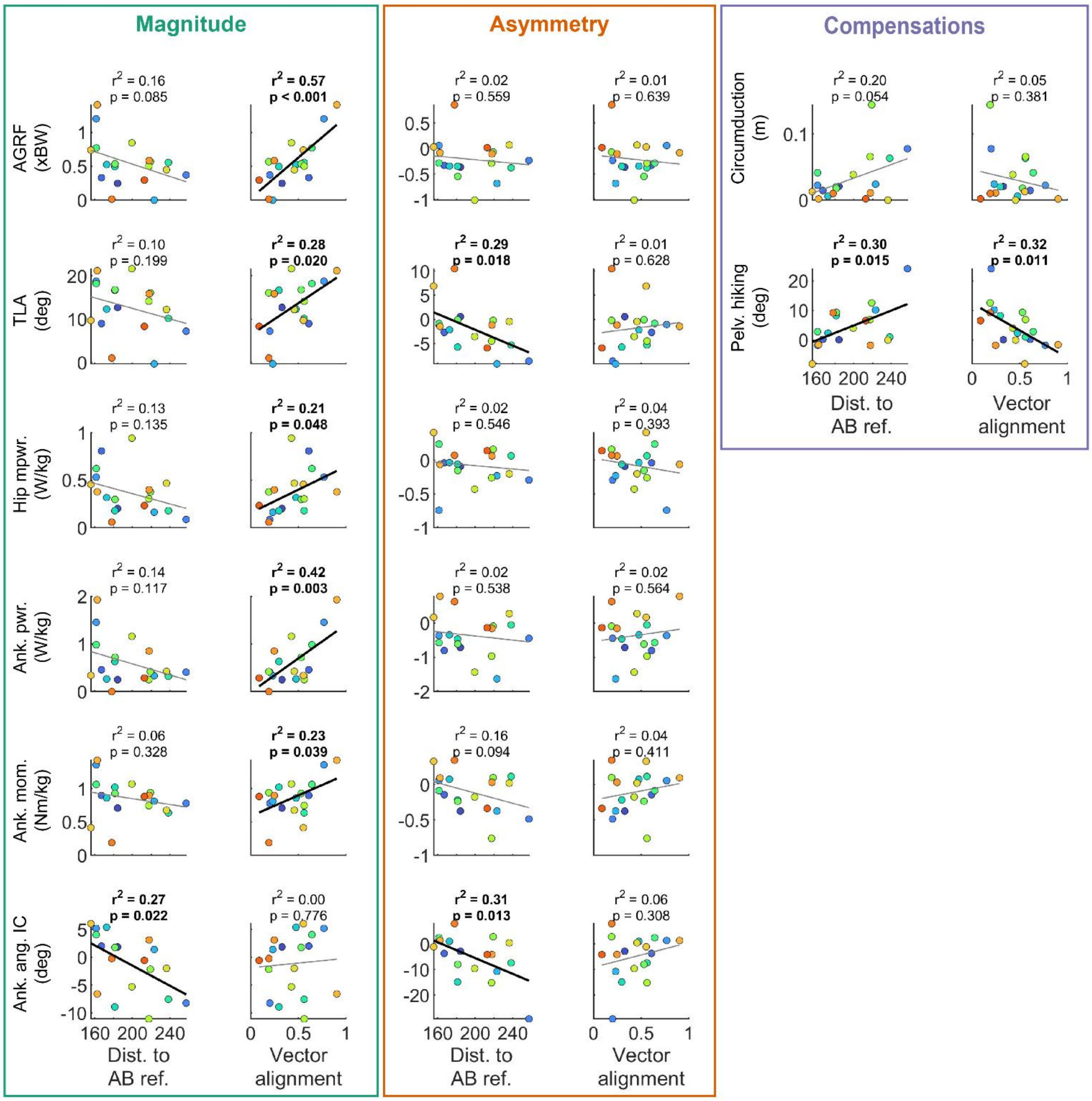


**Figure S4: Inter-individual correlations between post-stroke gait signatures and discrete biomechanical variables at participants' SS walking speeds**. Colors denote participants. Within each colored box, Euclidean distances to the AB reference gait signature (left column) and gait signature vector alignment (right column) correlations are shown side by side for each biomechanical variable. Left: Correlations with paretic-leg biomechanical magnitude variables (green box). Middle: Correlations with inter-limb asymmetry variables (orange box). Right: Correlations with paretic-leg compensation variables (purple box). The $r^2$ and corresponding p-values for a linear regression between one gait signatures metric and each biomechanical variable are shown in bold for $p < 0.05$. The thick black ($p < 0.05$) and thin gray ($p \geq 0.05$) lines show the linear regression estimate for each gait signatures metric and biomechanical variable.

Abbreviations: AGRF = anterior ground reaction force; Ank. Ang. IC = ankle angle at initial contact; Ank. mom. = peak ankle plantarflexor moment; Ank. pwr. = peak ankle plantarflexor power; Circ. = circumduction; Dist. To AB ref. = Euclidean distance to AB reference gait signature; Hip pwr. = Peak hip extensor power; Pelv. Hiking = pelvic hiking; TLA = trailing limb angle.

**Gait signature metrics regressed against changes in discrete biomechanical variables between the SS and fastest walking speeds.**

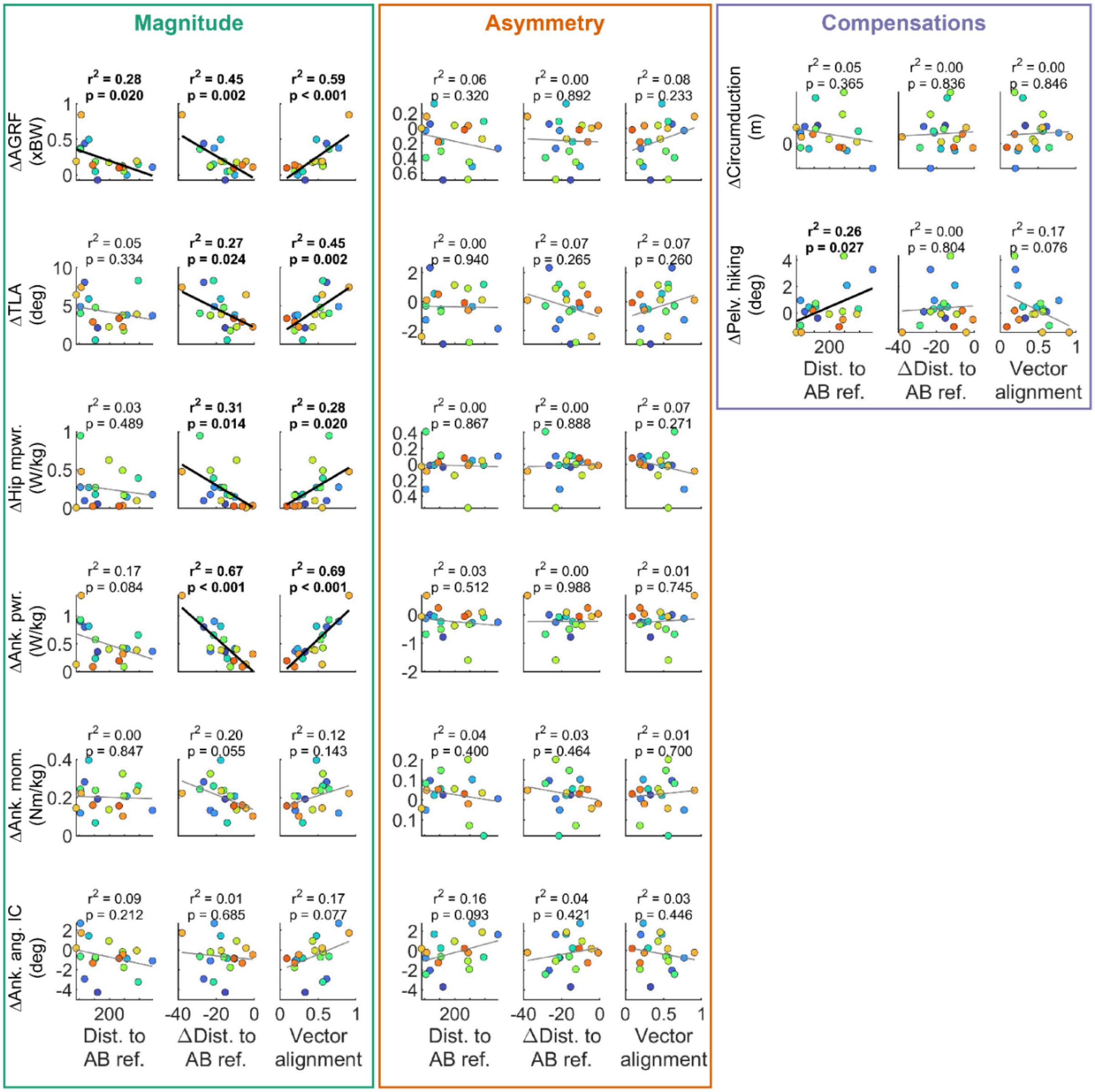


**Figure S5: Inter-individual correlations between post-stroke gait signatures and speed-induced changes in discrete biomechanical variables between participants' SS and fastest walking speeds**. Colors denote participants. Within each colored box, Euclidean distances to the AB reference gait signature (left column), speed-induced change in Euclidean distances to the AB reference (middle column; SS versus fastest speed), and gait signature vector alignment (right column) correlations are shown side by side for each biomechanical variable. Left: Correlations with paretic-leg biomechanical magnitude variables (green box). Middle: Correlations with inter-limb asymmetry variables (orange box). Right: Correlations with paretic-leg compensation variables (purple box). The $r^2$ and corresponding p-values for a linear regression between one gait signatures metric and each biomechanical variable are shown in bold for $p < 0.05$. The thick black ($p < 0.05$) and thin gray ($p \geq 0.05$) lines show the linear regression estimate for each gait signatures metric and biomechanical variable.

Abbreviations: AGRF = anterior ground reaction force; Ank. Ang. IC = ankle angle at initial contact; Ank. mom. = peak ankle plantarflexor moment; Ank. pwr. = peak ankle plantarflexor power; Circ. = circumduction; Euclidean distance to AB reference gait signature; Hip pwr. = Peak hip extensor power; Pelv. Hiking = pelvic hiking; TLA = trailing limb angle.
